# Microenvironment-driven phenotypic shifts in *Staphylococcus aureus* from children with cystic fibrosis: a longitudinal cohort study

**DOI:** 10.1101/2025.09.22.25336073

**Authors:** Stefania Robaldi, Rosana Pereda, Maria Constanza Pautasso, Enrique Blasko, Ailen Fretes, Trinidad Jaureguialzo, Mateo Diaz Appella, Mariana B. Galeano, Maria Jose Rial, Silvina Smith, Eugenia Ginestet, Norma Gonzalez, Javier Gonzalez, Andrea Smania, Alejandro Vila, Claudia Sola, Pablo M. Cassanelli, Paula M. Tribelli

## Abstract

*Staphylococcus aureus* (SA) is a common pathogen in children with cystic fibrosis (CF), frequently preceding or coexisting with *Pseudomonas aeruginosa* (PA). However, the extent to which its phenotypic traits are shaped by transient changes in the airway microenvironment, including coinfection and antibiotic exposure, remains poorly understood. In this study, we characterized 546 SA isolates from a longitudinal cohort of 16 pediatric CF patients over 18 months. Based on clinical records, patients were classified as SA-monoinfected or SA-PA-coinfected. Isolates were analyzed for staphyloxanthin production, hemolytic activity, DNase activity, biofilm formation, resistance to PA. Using principal component and hierarchical cluster analyses, we identified three phenotypic clusters. Isolates from monoinfected patients were predominantly found in a cluster characterized by high pigment and biofilm production and absence of chronic PA infection. In contrast, transient PA detection was associated with a shift toward a cluster displaying reduced pigment and biofilm levels. Additionally, in two out of five monoinfected patients, this shift coincided with ciprofloxacin eradication treatment including inhaled tobramycin or colistin and was accompanied by a significant increase in methicillin-resistant SA (MRSA) isolates. No phenotypic transitions were observed in chronically coinfected patients. Molecular characterization of a set of isolates showed on the one hand heterogeneity in the SA lineages and on the other hand the phenotypic profile was not determined by the genetic background. In vitro experiments showed a higher survival of MRSA over MSSA isolates in presence of both PA and ciprofloxacin. These results suggest that the phenotypic adaptation of SA in pediatric CF airway is driven by microenvironment rather than clonal background, and that transient coinfection and antibiotic exposure can favor MRSA emergence even in early-stage disease.

## Introduction

Cystic fibrosis (CF) stands as a frequent genetic disorder within the Caucasian demographic in which the cystic fibrosis transmembrane conductance regulator (CFTR) is mutated, impacting numerous organs and giving rise to diverse complications linked to patient mortality. Examples of such complications encompass cystic fibrosis liver disease (CFLD) and cystic fibrosis related diabetes (Malhotra et al.,2019). Chronic airway infections represent a leading cause of morbidity and mortality in these patients, with the burden being particularly severe in middle-and low-income countries due to limited access to specialized care and appropriate antimicrobial therapy (GBD 2021 Lower Respiratory Infections and Antimicrobial Resistance Collaborators). Although nowadays the use of cystic fibrosis modulators modified the CF scenario, bacterial infections and therefore its surveillance and treatment are still important due to the limitations of the modulator treatment such as age, CFTR mutation, patient nutritional condition and drug cost and accessibility (Guo et al., 2025).

In individuals with CF, the pulmonary microbiomes typically manifest dysbiosis alongside a pro-inflammatory milieu, leading to recurrent infections that may either be a consequence or a contributing factor, ultimately progressing to a chronic state over time (Dickison et al., 2021). *Staphylococcus aureus* is commonly acknowledged to be a causative agent during childhood, while *Pseudomonas aeruginosa* tends to be prevalent in patients as they age (Limoli et al., 2016; Malhotra et al., 2019). While this pattern holds true across various developed nations, it is crucial to recognize that infection dynamics may exhibit variations in middle or low-income countries.

Additionally, infections during childhood are important for CF patients since they provoke a progressive damage to lung tissue, decreasing the respiratory function and therefore impacting different aspects of their lives.

*S. aureus* and *P. aeruginosa* possess a wide metabolism and a set of virulence factors including quorum sensing molecules, biofilm formation, extracellular enzymes, and particularly *S. aureus* possess several toxins including hemolysins and immune system evasion proteins (Camus et al. 2021). The interaction between these bacterial species is complex, ranging from competition to coexistence and even metabolic cooperation. The main known functions that contribute to this interaction in-vitro have been revised recently by Biswas and Gotz (2022). These two bacterial species were isolated from patients, and their coexistence was also reported in vitro (Braiud et al., 2020). However, it is unclear how the presence of *P. aeruginosa* impacts on the selection of certain *S. aureus* phenotypes in vivo, particularly through time. To address these questions, we analyzed 546 *S. aureus* isolates obtained from a prospective and longitudinal study of 16 pediatric patients with CF diagnosis classified as *S. aureus* monoinfected or *S. aureus-P. aeruginosa* coinfected investigating the phenotype pattern of *S. aureus* isolates exposed to different airway microenvironments. In this work, we identified three major phenotypic clusters of *S. aureus* isolates (SA), independent of sequence type, based on virulence-and stress-related traits. Isolates from monoinfected patients predominantly belonged to a cluster defined by high staphyloxanthin and biofilm production. Transient detection of *P. aeruginosa* (PA) was associated with a phenotypic shift toward reduced pigment and biofilm levels, coinciding in some cases with eradication treatment with combined oral ciprofloxacin and inhaled tobramycin or colistin. In two patients, this transition was accompanied by a significant increase in MRSA frequency (P<0.001). These changes were not observed in patients with persistent PA coinfection, and along with a competitive advantage in vitro for a MRSA over a MSSA isolate in presence of both PA and a subinhibitory ciprofloxacin concentration highlighting the role of fluctuating environmental pressures in shaping SA phenotypes.

## Material and methods

### Clinical sampling design

Sixteen patients diagnosed with cystic fibrosis (CF) were randomly selected according to the following inclusion criteria. Patients were included if, at study initiation, at least two of their last four routine cultures were positive for *S. aureus* and they did not meet the criteria for chronic *P. aeruginosa* infection (hereafter referred to as mono-infected), or if they met the criteria for chronic infection with both microorganisms (co-infected). Chronic infection status was determined by the Bacteriology Department of the Central Laboratory at HGNPE. According to the Argentine Society of Pediatrics, chronic infection is defined as more than half of the respiratory cultures performed in the previous year being positive for a given microorganism, with a minimum of four routine cultures.

### Sampling

Isolates of *Staphylococcus aureus* (hereinafter SA) and *Pseudomonas aeruginosa* (hereinafter PA) were collected by the Bacteriology Department of the Central Laboratory at HGNPE from respiratory samples obtained during routine clinical visits or in the event of respiratory exacerbation over an 18-month period. For each respiratory sample collected, different colonies of *S. aureus* and *P. aeruginosa* were randomly selected (Fig. 1).

**Figure 1:**
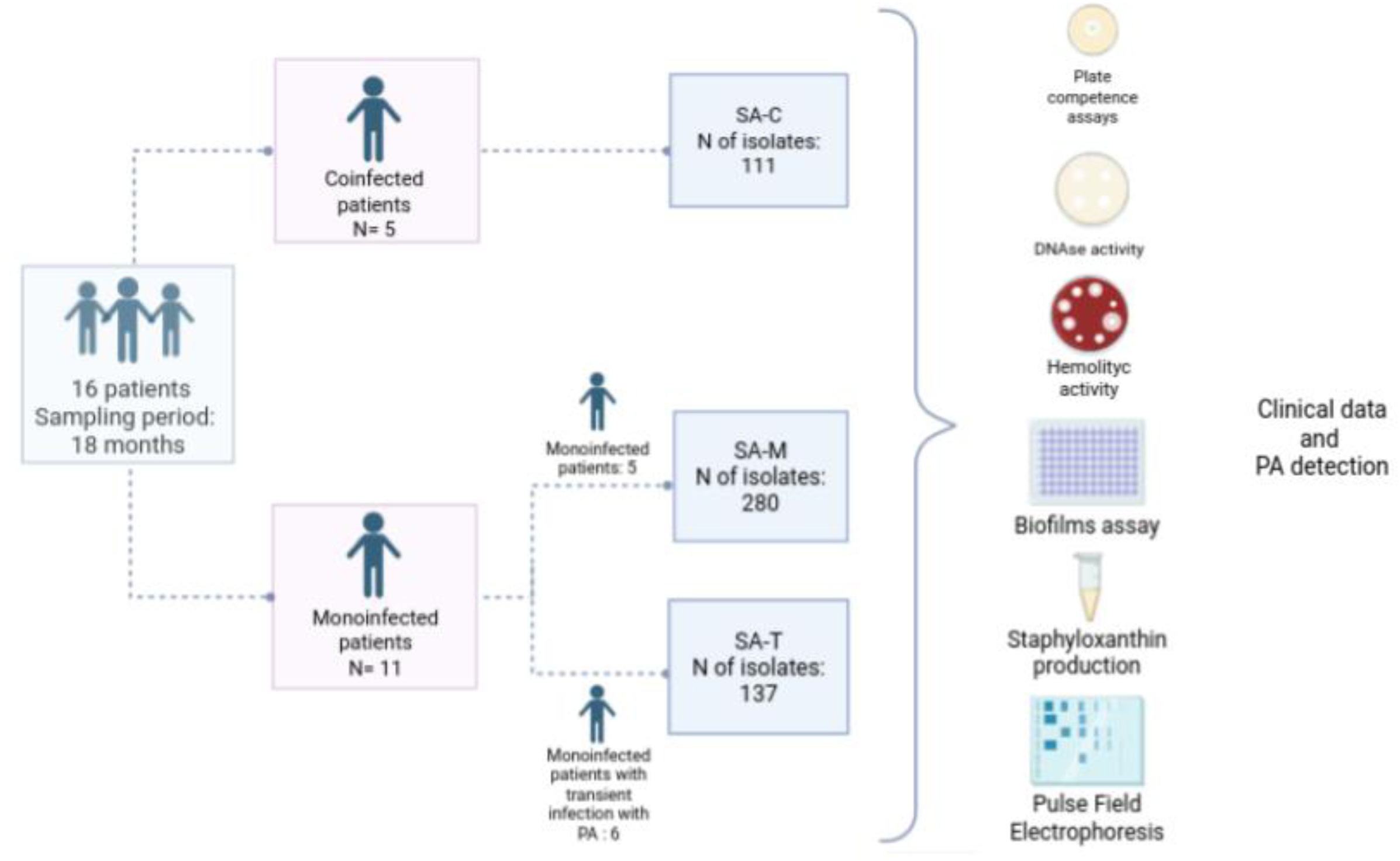
Clinical sampling design. Sixteen patients with CF diagnosis were classified as Mono-infected or Co-infected according to chronic infection status with SA or SA-PA. Isolates of SA and PA were collected from respiratory samples from these patients.

### Clinical Data Collection

Relevant clinical data generated during the sampling period were extracted from the digital clinical records of the recruited patients, including age, sex, CFTR variant, pulmonary exacerbations, antibiotic treatments, CFTR modulator treatment initiation and other pulmonary therapies. Furthermore, microorganisms isolated on routine and exacerbation cultures during this period, along with antibiotic susceptibility reports, were registered.

### Ethics statement

This study was reviewed and approved by the Ethical Review Board of General Children’s Hospital Dr. Pedro Elizalde (PRISBA 4466). All participants/patients (or their proxies/legal guardians) provided informed consent to participate in the study.

### Bacterial isolates and culture conditions

SA clinical isolates (N=546) were identified by standard microbiological procedures. SA cultures were grown in 5mL Tryptic soy Broth (Oxoid) (herein TSB) under aerobic conditions at 37°c overnight. Aliquots of these cultures were used for different phenotypic determinations

### Plate competence assays

Overnight cultures of SA isolates were adjusted to an optical density at 600nm (OD_600nm_) of 1 and seeded into Tryptic Soy Agar (TSA) plates. A 5-µL drop of a *P. aeruginosa* PAO1 culture, adjusted to an OD_600nm_of 10, was placed at the center of each plate. After drying, the plates were incubated overnight at 37°C. The diameter of growth inhibition zone and colony diameter were measured in each plate using ImageJ software.

### Virulence factors

For the quantification of the pigment staphyloxanthin, 1mL of the SA isolates cultures were centrifuged for 1 min at 13500 rpm and the supernatant was removed. Each pellet was resuspended in 1 mL of methanol and incubated at 37°C overnight in the dark with agitation. Tubes were centrifuged for 1 min at 13500 rpm and the supernatant was measured in spectrophotometer at 462 nm. A 5 µL drop of SA isolates cultures was placed in the corresponding plates and incubated at 37° C overnight. DNAse agar (Britania) was used to determine extracellular DNAse activity following manufacturer instructions. Hemolytic activity was analyzed in sheep blood agar 5%(v/v). Three biological replicates were measured for each isolate. For all tests, the degradation halo or clear zone was measured in each case, and the colony diameter was measured for normalization using the ImageJ software

### Biofilms assay

Biofilm formation was analyzed in 96-multiwell polystyrene plates using crystal violet assay. Briefly, SA isolates cultures were inoculated at an initial OD_600nm_ of 0.025 in TSB supplemented with 0.5% KNO_3_. The polystyrene plates were incubated overnight at 37°C. The supernatant was measured at 595 nm, and the adherent biomass was stained with crystal violet solution for 20 minutes. After incubation, the plate was washed, and the remaining dye was solubilized using absolute ethanol. The absorbance at 570nm was determined using BMG FLUOstar Optima Microplate Reader Fluorescence Absorbance Spectrometer. Biofilm index was calculated as the ratio between A_570nm_/A_595nm_.

### Molecular typing

Pulsed-field gel electrophoresis (PFGE) of SmaI-digested chromosomal DNA was performed on selected *S. aureus* isolates and interpreted as previously described (Barcudi 2024). *spa* typing was conducted on representative isolates from each PFGE-defined subtype as described previously, and spa types were assigned using the RIDOM web server (http://spaserver.ridom.de/). *spa* types were used to infer putative multilocus sequence types (MLST) based on established *spa*-MLST associations (Barcudi 2024, Barcudi 2020). When STs could not be determined from the *spa* server or from the literature, the genetic background of the isolates was determined by MLST as previously described (Barcudi 2024, Barcudi 2020 and reference therein). The specific sequence of the *sau1-hsdS1* gene characteristic of CC398, was also detected by PCR following the protocol of Stegger et al. (2011).

### Antibiotics sensitivity assays

Antibiotic sensitivity was analyzed in Mueller-Hinton agar (M-H, Oxoid) plates. Briefly, SA isolates cultures were adjusted to an OD_600nm_ of 0.5 and seeded into M-H plates. A disk of cefoxitin (FOX; 5 µg) was placed at the center of each plate. Plates were incubated at 37°C overnight. The diameter of the growth inhibition zone was measured in each plate using ImageJ software. Antibiotic susceptibility breakpoints were determined according to the Clinical and Laboratory Standards Institute (CLSI) guidelines (CLSI et al., 2023).

### Survival assays in presence of PA and ciprofloxacin

Minimum inhibitory concentration (MIC) assays for ciprofloxacin were performed following standard protocols. Briefly, serial two-fold dilutions of ciprofloxacin (Sigma) were prepared in sterile microplates using cation-adjusted Mueller–Hinton broth (Oxoid). Each well was inoculated with 1×10^5^ CFU of the corresponding *Staphylococcus aureus* isolate. Bacterial growth was monitored by measuring OD_600nm_every hour for 12 h at 37 °C under gentle agitation to minimize aggregation. One MRSA isolate and one MSSA isolate, both exhibiting a ciprofloxacin MIC of 0.5 µg/ml, were selected for subsequent experiments.

For coculture experiments, *Pseudomonas aeruginosa* PA14 and the selected *S. aureus* isolates (FO and EB) were pre-cultured on TSA prior to inoculation. Mono-and co-cultures were prepared as indicated in Table S1. Cultures were incubated on Muller-Hinton broth in microaerobic conditions at 37°C for 16 hours, and CFU/ml was determined by plating on Oxacillin Resistance Screening Agar Base (Oxoid) plates without oxacillin. The relative proportions of MRSA and MSSA colonies within cocultures were further assessed by replica plating onto oxacillin (4 µg/ml) (Sigma) containing agar plates.

### Data analysis

Continuous variables were expressed as median and interquartile range (IQR). Association between phenotypes and chronic PA infection status was performed using Mann-Whitney’s test. Correlations between phenotypes were evaluated with Spearman’s rank test. In addition, principal component analysis of the phenotypes was carried out using the *FactoMineR* package (Lê et al.,2008). The principal components obtained were then used to perform hierarchical clustering analysis with Ward’s method (Ward et al., 1968). Association between the clustering and clinical data, such as patient, chronic PA infection, modulator treatment or co-isolation with *Candida* spp. or *Aspergillus* spp., was then analyzed using Fisher’s exact test. Proportions of MRSA of different sampling dates were compared using Fisher’s exact test. All statistical analyses were performed with GRAPH-PAD PRISM version 8.0.2 (Dotmatics, Boston, MA, USA) for Windows and R version 4.4.2. and a P<0.05 was considered as statistically significant.

## Results

### Patient Clinical Profiles

The clinical and microbiological characteristics of the cohort consisting of 16 pediatric patients are summarized in Table S2 and the collection scheme in Fig 1. The median age at the onset of the sampling period was 8.8 years old (IQR 6.5 - 13.2 yo) with different severity in VEF affectation according to the criteria used in the Cystic Fibrosis Foundation annual report. The most frequent variant was F508del with 8 heterozygous and 5 homozygous patients. Five patients carried additional variants to F508del. One patient was diagnosed using clinical criteria (positive newborn screening and abnormal sweat chloride) but had no CF-associated variants detected. CFTR modulator therapy was introduced in 2020, and this work included 2 patients already receiving the combination lumacaftor/ivacaftor at the beginning of the study, 8 patients who began treatment at some point during the sampling period and 6 patients not treated with modulators (Table S2). The annual median of exacerbation in this cohort of patients was 3.8 episodes and antibiotic courses, inhaled DNAse and other treatments are summarized in Table S2.

All recruited patients were chronically infected with *Staphylococcus aureus* (SA) or co-infected with *Staphylococcus aureus* and *Pseudomonas aeruginosa* (PA) at the start of the sampling period, resulting in 10 SA-monoinfected and 6 SA-PA-coinfected patients. The number of SA isolates from each patient is shown in Table S2 with a median of 30.5 (IQR: 20.5-44.5).

### Phenotypical characterization of *S. aureus* isolates from pediatric cystic fibrosis patients

We analyzed a total of 546 SA isolates, collected from 11 patients classified as SA monoinfected and 5 classified as SA-PA-coinfected. The sampling included isolates from routine cultures and cultures requested during exacerbations performed between July 2021 and January 2023 (Fig. 1). Given the dynamic nature of infections and the sampling period, SA isolates were classified as follows: SA-M, isolates from patients recruited as monoinfected who did not develop any PA infection during the sampling period, or collected before a PA transient infection; SA-T, isolates obtained from monoinfected patients during or after a transient PA infection; and SA-C, isolates from patients recruited as coinfected (Fig. 1).

We selected phenotypes related to virulence and stress resistance: DNAse and hemolytic activity as well as staphyloxanthin production and biofilm formation. Agar plate competence assays using *P. aeruginosa* PAO1 against the different SA isolates from the patients were also performed. These phenotypic features were assayed for 546 isolates and were analyzed using different strategies of multivariate statistical analysis to understand the relationship between virulence and infection state through time.

As a first approach we analyzed the different phenotypes individually using the SA mentioned classification. The median of competence and biofilm did not show any differences among groups (Fig. 2 A and B). Hemolytic activity showed differences among groups, but mostly due to the presence of non-hemolytic isolates, therefore a Chi Square-test was conducted showing differences between the SA-M and SA–C group with higher proportion in SA-M ( ^2^= 3.860, df=1, p value = 0.0494, 16.78% in SA-M vs 9.01% in SA-C) (Fig. 2C and D). Staphyloxanthin content differed among groups being the lowest in the SA-C (SA-C mean= 0.1521 n= 111, SA-M mean= 0.1886 n=280 and SA-T mean= 0.2452 n=137) (Fig. 2E). Extracellular DNAase activity showed differences between groups, being the higher value for SA-M group and the lowest for SA-C group (Fig. 2B).

**Figure 2:**
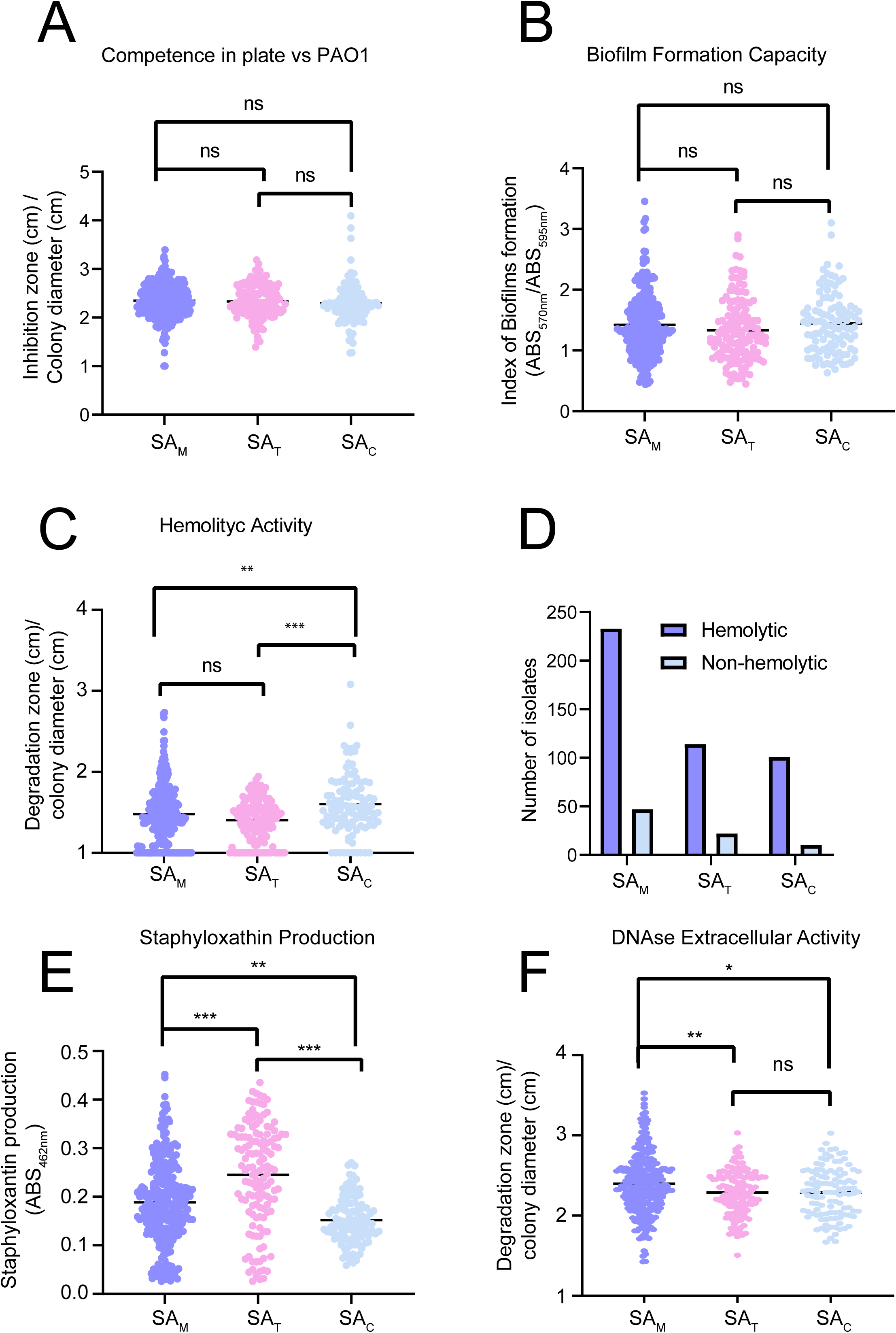
Phenotypes of SA isolates. A. Competence against *P. aeruginosa* PAO1 reference strain. B. DNAse activity in DNA-agar plates. C. Hemolytic activity in sheep agar blood. D. Non-hemolytic isolates proportion. E. Staphyloxanthin production obtained after methanol extraction F. Biofilm formation in polystyrene microplates determined by Crystal Violet stain. One-way Anova was used (* P 0.05; ** P 0.005 and *** P 0.0005).

We then assessed Spearman’s rank correlation between the phenotypes. Both biofilm formation and staphyloxanthin were found to negatively correlate with DNAse activity (rho =-0.23, p < 0.001; rho =-0.15, p < 0.001, respectively), while hemolysis correlated positively with the latter (rho = 0.13, p < 0.01) and with competence (rho = 0.13, p < 0.01). These correlations, although weak, suggest that some aspects of the underlying regulation of these traits could be shared (Table S3).

We performed PCA on all phenotypes from the 546 SA isolates (Fig.S1). Components 1 and 2 explained 51.11% of the variance (27.51% and 23.60%, respectively). Biofilm formation and DNAse activity were the main contributors to component 1 (r =-0.723 and r = 0.628), while staphyloxanthin, DNAse, and competence correlated with component 2 (r = 0.758, r =-0.485, and r = 0.54, respectively). Variable vectors for components 1 and 3 accounted for 46.38% of the variance, with component 3 explaining 18.87% (Fig. S1). Hemolysis correlated positively with component 3 (r = 0.82; Table S4).

Afterwards, hierarchical clustering analysis was conducted based on the principal component analysis (PCA). As is shown in Fig. 3 A and B, SA isolates could be classified in three defined clusters named from Cluster 1 to 3. Each cluster showed a similar number of assigned isolates with 199 isolates classified as Cluster 1, 139 as Cluster 2 and 188 as Cluster 3 and all of them presented contributions of different patients (Fig. S2 A). Cluster 1 was characterized by high staphyloxanthin and biofilm production. Additionally, the competence was higher in comparison with other groups (Fig. 3A). The most relevant characteristic of Cluster 2 was the low hemolytic activity and low staphyloxanthin production (Fig. 3A). Cluster 3 isolates showed low biofilm formation and low competence with PA as well as high DNAse production (Fig. 3A). The median of each group for each phenotype was summarized in Fig. 3B.

**Figure 3:**
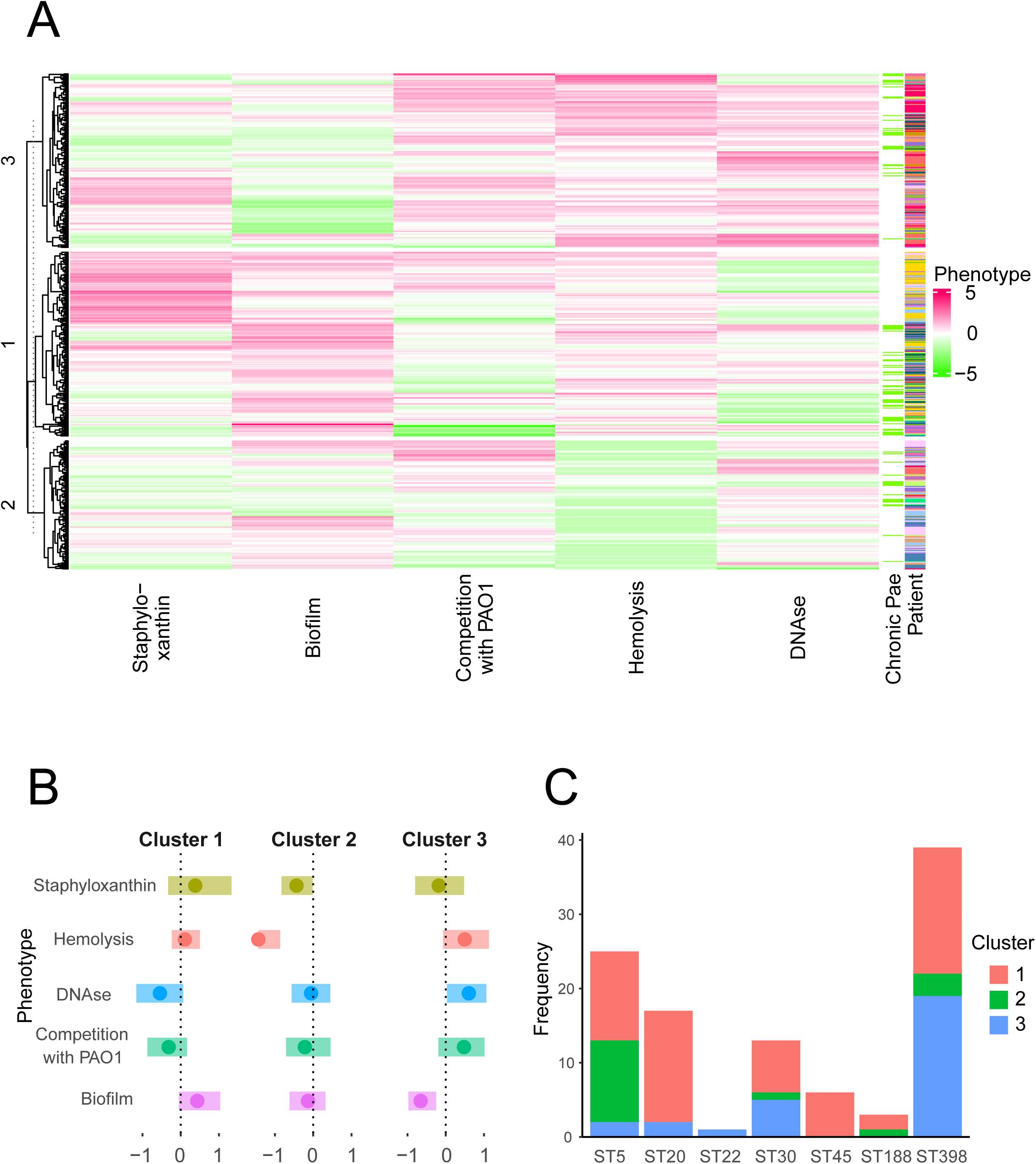
Phenotypic clustering of SA isolates A. Hierarchical clustering analysis. Different phenotypes, presence of chronic PA and patient origin are shown. Colors indicate the median of the individual isolate. B. Representation of phenotypic profile for each cluster. The zero represents the mediane of a certain phenotype calculated with the 546 analyzed isolates.

A further analysis between clusters and SA groups (M, T or C), although association was not significant, showed a trend where Cluster 1 is associated mainly with SA-C while Cluster 3 is associated mainly with SA-M and SA-T (Fig. S2 B). We sought to identify the factors that determine the phenotypic values underlying the observed clustering. To this end, we analyzed the association of each cluster (in terms of the number of isolates) with the SA group (M, T or C), the use of CFTR modulators, the presence of chronic PA infection, and the individual patient. Our results showed that Cluster 3 had an enrichment in SA isolates from patients using the modulator therapy at any point of the sampling period (Fisher Test, p<0.01; Fig. S2C) while the presence of chronic PA did not show any association with a specific cluster (Fig. S2D). However, an absence of chronic PA can be observed in a defined group of Cluster 1 with high staphyloxanthin production (Fig. 3A).

Additionally, we questioned whether the clusters were composed of specific *S. aureus* lineages or clonal complexes (CCs). To investigate this, we selected isolates from different clusters and performed pulsed-field gel electrophoresis (PFGE) complementing in selective isolates with molecular *spa* or MLST typing (Supplementary Fig. S3A and B; Table S5). Our results primarily showed a diversity of *S. aureus* lineages within a cluster, including ST398 (CC398), ST5, ST20, ST30 (CC30), ST22 (CC22), ST45 (CC45), and ST188 (CC1), highlighting the genetic variability among patients and even within individual patients. MSSA CC398 appeared to be the most frequent lineage among the recruited patients (Table S5). PFGE profiles within individual patients were variable; for example, patient 1 (hereinafter P1) consistently showed ST20 isolates with the same *spa* type across samples, until transient PA infection where we found PFGE and *spa* variations corresponding with ST30 strains. In contrast, other patients such as P9 and P18 exhibited different STs—ST45 and ST398 in P9, and ST5 and ST398 in P18. Isolates with identical *spa* and ST showed different phenotypic profiles across patients. For instance, in P12, P9, and P18, isolates with *spa* 1451/ST398 fell into different clusters depending on the patient. The same occurred for P1, P2, and P9, where isolates with the same molecular type were assigned to distinct phenotypic clusters (Table S5).

Our results showed that phenotypic clustering was not determined by the sequence type (ST), as different STs were distributed across distinct clusters (Fig. 3C). Similar findings were observed when individual phenotypes were analyzed, indicating that specific phenotypic values were not associated with a particular lineage.

### SA phenotypes are affected by airway microenvironment

Infection in children with CF is dynamic and could change over time. For example, patients recruited as coinfected may have initiated triple therapy with elexacaftor/tezacaftor/ivacaftor (ETI) during the sampling period, or an SA-monoinfected patient may have experienced a transitory PA infection (Table S2). Therefore, we decided to analyze the SA isolates, based on the five measured phenotypes that define the clusters, from each patient over time, focusing on PA decrease or transient infection; the presence of *Candida* spp. or *Aspergillus*; initiation of modulator therapy; and antibiotic treatment, to understand SA phenotype dynamics. We selected patients which were representative of different scenarios.

First, we analyzed monoinfected patients P3, P11 and P12 which showed all SA-M since no PA was isolated during the sampling time (Fig.1). In P3, a stable Cluster 3 pattern was observed, except for a single time point in May 2022, which coincided with empiric ciprofloxacin treatment due to a worsening of clinical parameters, although no Gram-negative bacteria were isolated (Fig 4A). P11, is a child who underwent changes in modulator therapy. At the time of recruitment, the patient was receiving lumacaftor/ivacaftor, which was replaced by ETI in January 2022. Five months later, SA isolates displayed variable phenotypes, with a shift from Cluster 2 to Cluster 3, which was maintained in subsequent samplings (Fig. 4B). In P12, isolates mainly belonged to Cluster 3, characterized by low staphyloxanthin production, high DNase activity, and variable biofilm formation and hemolysis. This patient underwent antibiotic treatment around August 2022 due to lung function deterioration—initially with oral ciprofloxacin and trimethoprim-sulfamethoxazole (TMS), followed by two hospitalizations requiring intravenous amikacin and vancomycin. The isolates obtained during and after these antibiotic treatments exhibited phenotypic changes, including decreased hemolysis and biofilm formation (Fig. 4C).

**Figure 4:**
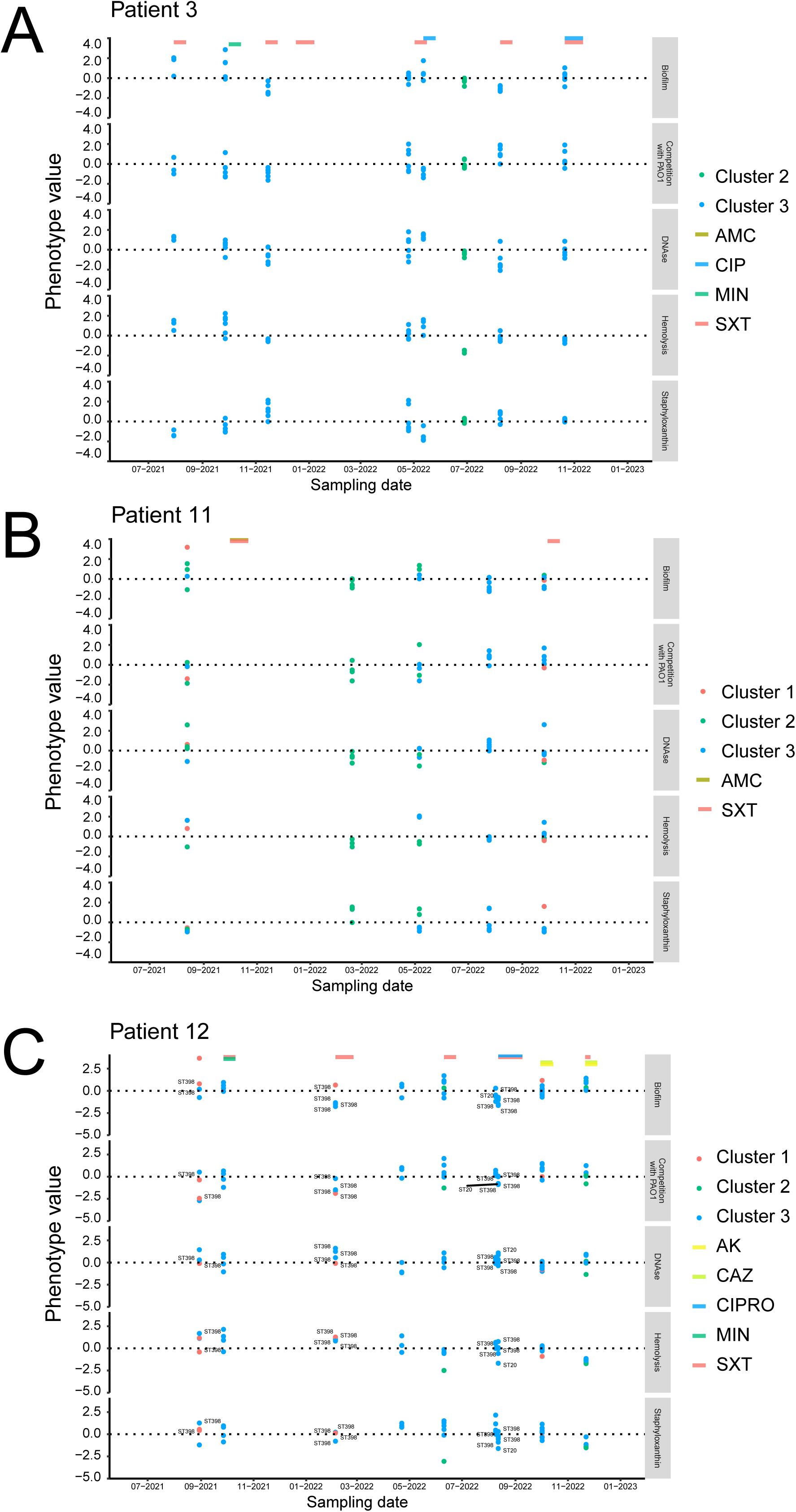
Temporal dynamics of the phenotype values in SA isolates from P3, P11 and P12 all displaying SA-M. Each data point represents the normalized mean value from three independent determinations of biofilm formation, competition with PAO1, DNAse activity, hemolysis or staphyloxanthin production for a single isolate. At each sampling time point, multiple isolates were randomly selected from patients undergoing routine clinical monitoring or experiencing exacerbations.

We also analyzed three patients who were monoinfected that underwent transitory PA infections, thus we obtained SA-M (before the PA infection) and SA-T isolates during and after the PA infection (Fig.1). In P1, most SA-M isolates belonged to Cluster 1, characterized by high staphyloxanthin production and DNase activity while hemolysis and biofilm formation were low (Fig. 5A). However, when a transient PA infection occurred, along with oral ciprofloxacin treatment combined with inhaled colistin, SA-T isolates shifted to Cluster 3, exhibiting lower values in biofilm formation and staphyloxanthin production, while the other parameters remained largely unchanged (Fig. 5A). P2 suffered a transitory PA infection along with ciprofloxacin treatment in combination with inhaled tobramycin with an increase in SA-T isolates belonging to Cluster 2 with staphyloxanthin production, hemolysis and biofilm formation altered while the rest of the parameters were similar (Fig 5B). DNAse treatment started in this patient in 2022-September and after this month isolates were all grouped in Cluster 3 (Fig 5B). P15 is particularly interesting, as this child was recruited as monoinfected but experienced a nearly year-long PA infection. During the initial period, SA-M isolates belonged to Cluster 1, and after approximately four months, the SA-T isolates were classified Cluster 2 exhibiting decreased staphyloxanthin production, biofilm formation and hemolytic activity while DNase activity was increased (Fig. 5C). Additionally, this patient initiated ETI in June 2022. While PA was no longer isolated, SA continued being recovered, displaying a progressive decrease in hemolysis even after PA clearance, whereas biofilm formation and staphyloxanthin levels were restored to earlier values (Fig. 5C).

**Figure 5:**
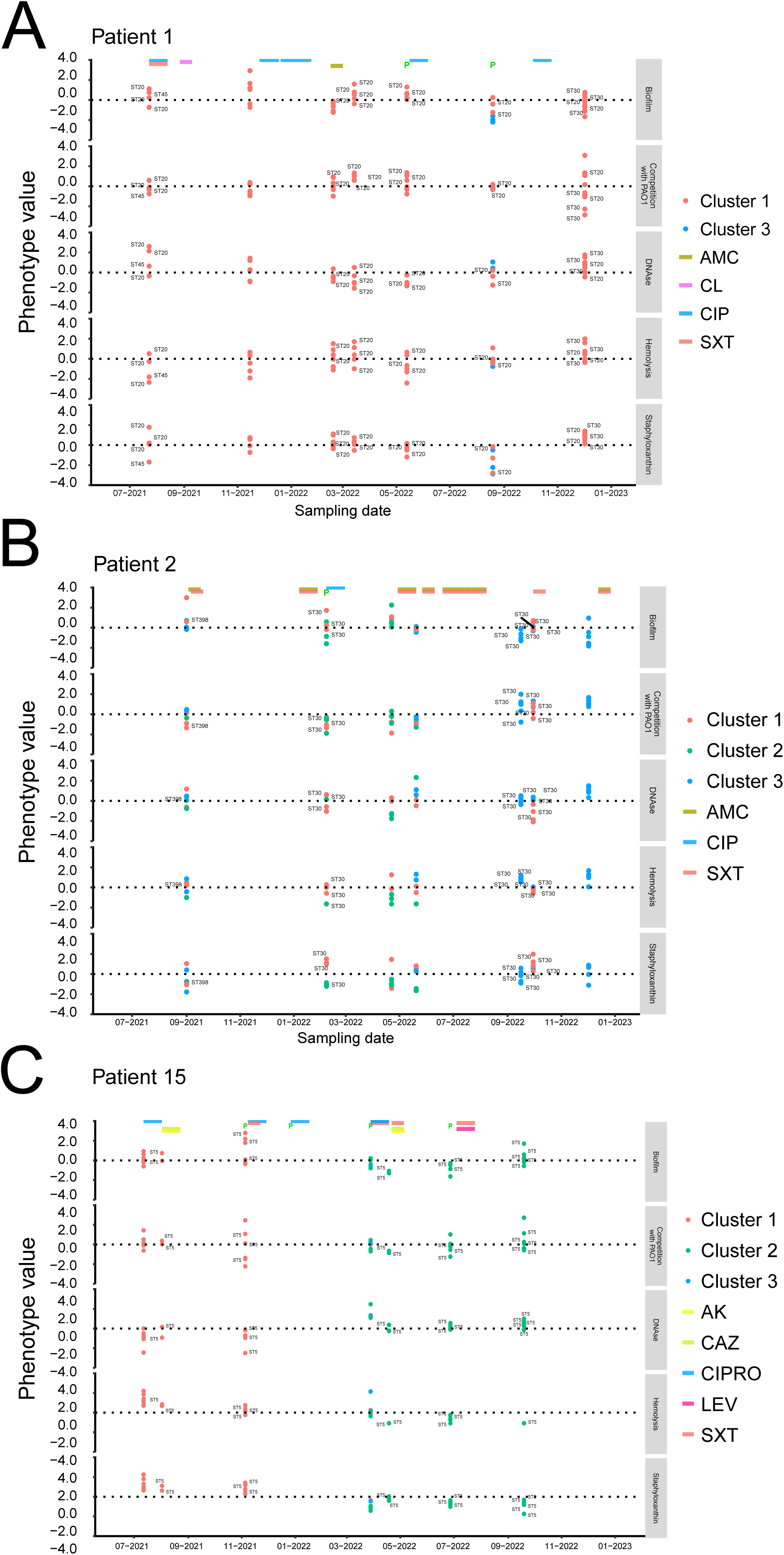
Temporal dynamics of the phenotype values in SA isolates from P1, P2 and P15 all displaying SA-M and SA-T. Each data point represents the normalized mean value from three independent determinations of biofilm formation, competition with PAO1, DNAse activity, hemolysis or staphyloxanthin production for a single isolate. At each sampling time point, multiple isolates were randomly selected from patients undergoing routine clinical monitoring or experiencing exacerbations.

Finally, for coinfected patients, we selected P7, P8 and P9 for longitudinal analysis with all SA-C isolates. P7 began ETI during the study period, being exposed to the treatment after April 2022. Prior to modulator therapy, SA-C isolates from this patient were grouped into Clusters 1 and 3, with Cluster 1 being the most frequent. After April 2022, PA was no longer isolated, while SA isolates were predominantly assigned to Cluster 3, displaying altered hemolytic activity and more variable biofilm formation (Fig. 6A). For P8 and P9 we observed a different phenomenon probably due to SA displacement or sub diagnosis provoked by PA presence. Sampling from P8 was characterized by the absence of SA for several months. When SA isolates reappeared, a mixture of isolates belonging to the three clusters was observed. However, less than two months later, all the SA isolated were classified as Cluster 2 by our algorithm (Fig 6 B and C).

**Figure 6:**
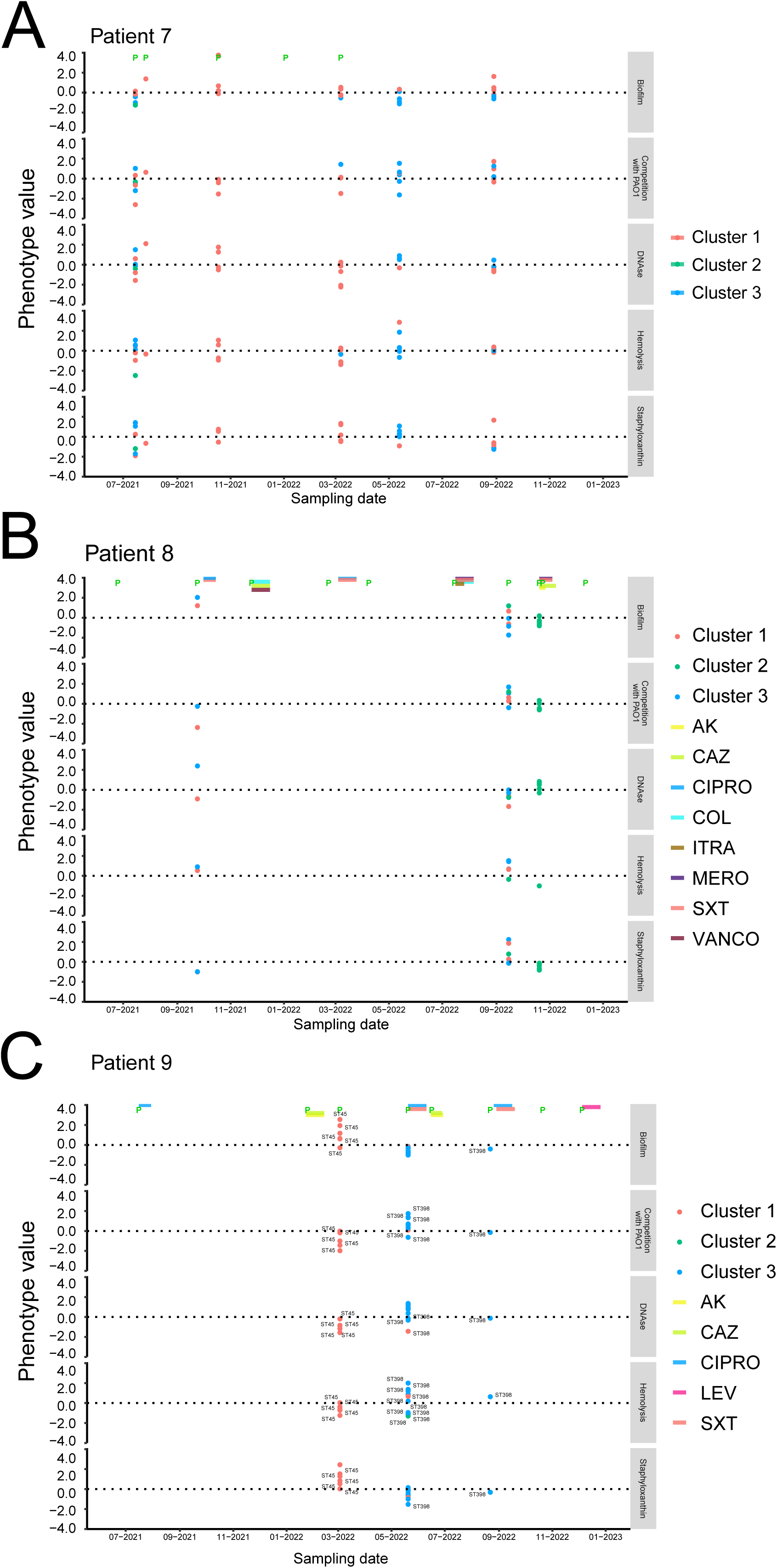
Temporal dynamics of the phenotype values in SA isolates from P7, P8 and P9 all displaying SA-C. Each data point represents the normalized mean value from three independent determinations of biofilm formation, competition with PAO1, DNAse activity, hemolysis or staphyloxanthin production for a single isolate. At each sampling time point, multiple isolates were randomly selected from patients undergoing routine clinical monitoring or experiencing exacerbations.

Overall, our results showed that over time, SA phenotype clustering varied following fluctuations in PA presence, initiation of modulator therapy, or antibiotic treatment targeting Gram-negative bacteria such as ciprofloxacin. These changes were not associated with the SA clonal complex, but rather with the surrounding microenvironment, highlighting the importance of monitoring SA infections following treatment modifications or changes in patient routine (Fig 4-6 and Fig. S4-10).

### Transitory *P. aeruginosa* infection along with Ciprofloxacin treatment select MRSA over MSSA

As MRSA represents a clinical threat in children with CF, we analyzed methicillin resistance in different patients, particularly those who experienced transient *P. aeruginosa* infection treated with ciprofloxacin. Methicillin resistance was initially reported by the hospital’s clinical microbiology service using standard clinical methods while we analyzed the individual isolates. For P1 and P2 the bacteriology department reported a change from MSSA to MRSA infection. When we analyzed this change, we found out that it was concomitant or immediately after *P. aeruginosa* isolation along with ciprofloxacin treatment. We therefore tested the proportion of methicillin resistant isolates (using resistance to FOX as a surrogate marker following CLSI guidelines) comparing SA-M and SA-T of the following patients: P1, P2, P4, P5 and P15 (Table 1). In P4 and P15 no isolates presented methicillin resistance while in P5 the proportion of MRSA was low and similar for SA-M and SA-T groups (Table 1). For P1 and P2, a Fisher’s exact test revealed a significant increase in the percentage of MRSA isolates, rising from 0% to 14.6% and 0% to 77.8% respectively after *P. aeruginosa* infection and ciprofloxacin treatment consistent with clinical records (Table 1, Fisher Test p<0.001). Interestingly, P1 and P2’s age was below 6 years old, and no modulator therapy was used for their cases. On the other hand, P5 and P15 used CF modulators and these patients with the addition of P4, were between 9-14 years old. The change of MSSA population to a MRSA lineage was not observed for chronic PA infection despite the different applied therapies.

**Table 1.** MRSA and MSSA proportion in SA-M and SA-T from patients with Pae transitory infection and treated with ciprofloxacin.

| Patient | Number of isolates | % of isolates |  |  |  |
| --- | --- | --- | --- | --- | --- |
|  |  | SA-M<br>MRSA | SA-T<br>MRSA | SA-M<br>MSSA | SA-T<br>MSSA |
| Patient 1 | 48 | 0 | 14.6 | 37.5 | 47.9 |
| Patient 2 | 45 | 0 | 77.8 | 13.3 | 8.9 |
| Patient 4 | 29 | 0 | 0 | 0 | 100 |
| Patient 5 | 18 | 16.7 | 5.6 | 5.6 | 72.3 |
| Patient 15 | 35 | 0 | 0 | 22.9 | 77.3 |

We therefore hypothesized that *P. aeruginosa* involved in transient infections may exhibit environmental-like traits—such as LasA and pyocyanin production—that, in combination with ciprofloxacin, selectively impair MSSA and thereby favor MRSA persistence. We employed the *P. aeruginosa* PA14 strain (PA14), a highly virulent isolate with robust pyocyanin production, resembling environmental or transient *P. aeruginosa*. To select ciprofloxacin-resistant *S. aureus* isolates for *in vitro* experiments, we first performed standard MIC assays. One MRSA and one MSSA isolate, both from P2 that showed a 0.50 µg/ml MIC of ciprofloxacin, were selected and incubated either alone or in co-culture with PA14, in the presence or absence of a subinhibitory concentration of ciprofloxacin (0.25 µg/ml) (Table S1). MRSA and MSSA showed comparable CFU/ml in control monocultures and when exposed to PA14 or ciprofloxacin individually. However, in co-cultures containing MRSA + MSSA + PA14 in ciprofloxacin supplemented cultures, we observed a marked increase in the proportion of MRSA, an effect not detected in the absence of the antibiotic (Fig. 7).

**Figure 7:**
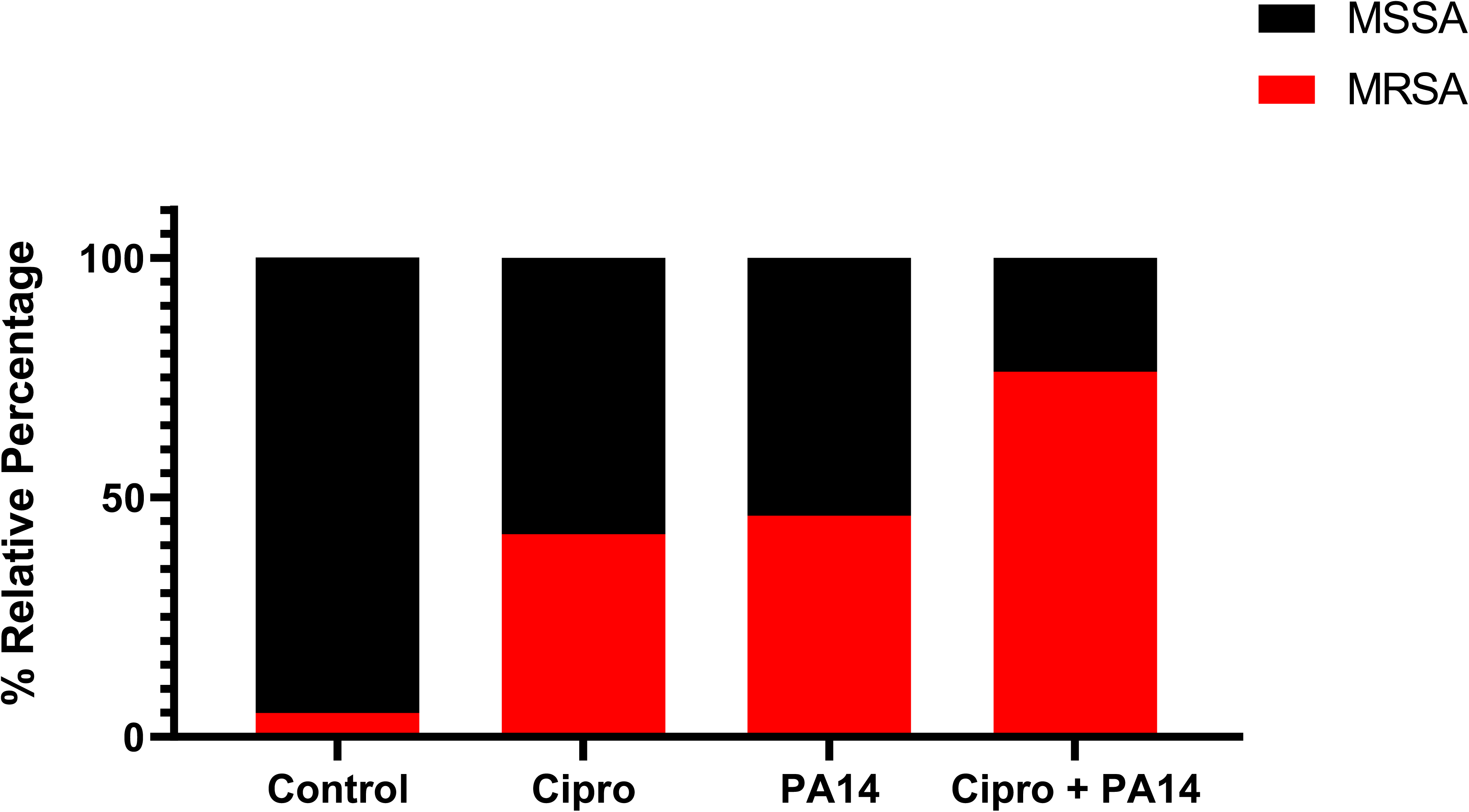
Impact of PA14 in MSSA and MRSA isolates survival in presence and absence of ciprofloxacin. The % of MRSA or MSSA after cultivation was determined by plating followed by replica plating in oxacillin plates.

## Discussion

Polymicrobial infections are complexly affected by a variety of factors, including the pathological scenario, the immune system of the host, the genetic background of the circulating strains, the native microbiome but also the antibiotic and other drug therapies as well as the presence of different transient or chronic, opportunistic or pathogenic bacterial or fungal species (Reece et al., 2021). In several complex clinical scenarios polymicrobial infections lead to a worse patient prognosis, for example in diabetic foot, burn wounds, catheter-associated infections, brain abscesses and pelvic inflammatory disease, among others (Eichorn et al., 2024; Haggerty et al. 2016; Barshes et al., 2022; Sen P et al., 2020; Roy et al., 2022; Nye et al., 2024). Additionally, primary ciliary dyskinesia and cystic fibrosis are conditions where a chronic polymicrobial airway infection is established (Wijers et al., 2017). Cystic Fibrosis (or CF) is a genetic disease characterized by an inflammatory airway environment and bronchiectasis that has the need for a lung transplant or the death of the patient as a final consequence. CF has been the hallmark of chronic polymicrobial infections with an initial colonization of *Haemophilus influenzae* and *S. aureus* during childhood while a posterior *P. aeruginosa* infection is frequently found with the establishment of a coinfection or *S. aureus* displacement (Budden et al., 2019).

However, differences in the colonization dynamics can be observed in different countries depending on the access to health care and early diagnosis, for example Rosales-Reyes et al., (2020) reported that the 47.5% of patients between 0-18 years old were colonized by *P. aeruginosa* while in with a life expectancy of 19 years. In European countries life expectancy reaches to a median of 51.3 and the difference among countries is mainly due to socioeconomic factors (McKone et a., 2021).

Nowadays CF modulator therapy has changed the life perspective for the patients, and gene editing is now possible, but there are still limitations related to CFTR mutations, patient age, metabolic patient profile and economical access to the treatment (Jia et al., 2023). Besides the improvement of lung function and the increase in life expectancy, the presence of pathogenic bacterial species like *S. aureus* and *P. aeruginosa* is still a concern (Milczewska et al.,2024). In Argentina, the access to the CF modulator was first available in 2020 but after October 2021 a local company started to produce the CF modulators allowing a better access to it. However, the healthcare system includes a complex bureaucratic pathway that can complicate the starting and maintenance of the treatment. In this work we analyzed 16 pediatric patients from a hospital in Buenos Aires, Argentina with different CFTR mutations and a variety of clinical profiles, including mild, moderate and severe lung function impairment. Additionally, they presented different microbiological status, that is young children where a recent staphylococcal chronic infection was acquired alongside with older children and adolescents with either a dominant SA chronic infection or chronic SA-PA coinfection with episodic isolation of fungal species. These patients had access to multidisciplinary follow-up, pulmonary therapies (inhaled hypertonic saline and DNAse, macrolides, inhaled antibiotics, supplementary oxygen and non-invasive ventilation) and complementary diagnostic exams when required. CFTR modulator therapy was requested by the patients to health providers, which responded favorably although with some delay.

Bacterial interactions are complex and different models that explain these interactions have been recently revised by Srinivasan et al., (2024), including those observed for *P. aeruginosa* and *S. aureus*. These bacterial species display different physiological responses ranging from antagonism to metabolic syntropy in CF patients mediated by the acetoin metabolism and cooperation in wounds through a division of labor (Camus et al., 2020). Additionally, more complicated scenarios are observed if the initial colonizer is considered in *in vitro* experiments since in the patients the presence of *S. aureus* in the skin could lead to an initial advantage to colonize the host and the cost of infection for *P. aeruginosa* increases with the concomitant increment of virulence factors to cope with the competitor (Srinivasan et al., 2024). Here we followed the *S. aureus* infection dynamics of CF pediatric patients classified as monoinfected with SA (mono) or coinfected with SA-PA (co). Using five different phenotypes (biofilm formation, staphyloxanthin production, competence against *P. aeruginosa* PAO1 and DNAse and hemolytic activity) a clustering analysis allowed us to group the SA isolates in three main groups displaying a different phenotypic pattern. Although the experimental design limitations we analyzed the factors that could drive this clustering and in some point to predict the infection dynamics.

Our results showed that the *S. aureus* isolates were grouped in three clusters in which changes in hemolysis, biofilm and staphyloxanthin production seem to be relevant factors in determining regrouping after disturbing events such as PA transient infection or CF modulator therapy initialization although we cannot rule out other external or patient related factors. Particularly, we observed a change to a pattern with lower biofilm formation and staphyloxanthin production after PA transitory infection.

Staphyloxanthin is a triterpenoid that is embedded in the cell membrane, and its biosynthesis depends on the presence of the *crtOPQMN* operon driven by a single promoter. Contradictory results can be found in literature regarding pigment production, while Antonic et al., (2013) reported that staphyloxanthin production is increased in presence of *P. aeruginosa* since this pigment protect *S. aureus* from pyocyanin derived oxidative stress, Biswas et al.,(2009) reported that the presence of *P. aeruginosa* lead to small colony variant phenotype with low staphyloxanthin production showing the complex interaction between these species. We found that staphyloxanthin production was higher in isolates belonging to monoinfected patients compared with those isolated from chronic coinfected patients. Besides pigment production, we analyzed biofilm formation in *S. aureus* that depends on the exopolysaccharide production derived from the *ica* operon but also the contribution of proteins and adhesins (Fbn, Clumping factors and Spa). Interestingly, *S. aureus* biofilm formation as well as staphyloxanthin production is regulated positively by SarA and SigB regulators. A recent study showed that clinical *S. aureus* cocultured with clinical *P. aeruginosa in vitro* showed a sharp decrease in the expression of *spaA*, *cflA* and *icaA,* all of which are involved in biofilm formation, compared with cocultures with the reference strain PAO1 while *sarA* expression was found to be unchanged with respect to *S. aureus* monocultures (Wang et al., 2025). Briaud et al., (2019) analyzed the coexistence or competence of clinical isolates from chronically coinfected CF patients showing a repression of-2.49 folds of *sarS* in a coexistence with a clinical isolate of *P. aeruginosa*. Interestingly, Tognon et al., (2019) analyzed the gene expression profile of *S. aureus* after 3h of coculture with *P. aeruginosa* and found that *sarA* and *sarR* were repressed in cocultures compared to monocultures (Tognon et al., 2019). All these reports showed the complex *S. aureus* response to *P. aeruginosa* and the technical limitation to study a large number of *S. aureus* in vivo complicates a complete interpretation. However, our work analyzed more than 500 *S. aureus* isolates from 16 CF pediatric patients showing a clear impact in the *S. aureus* phenotype when these isolates are exposed to *P. aeruginosa*, antibiotic or CF therapy. Although the clustering of *S. aureus* coexisting chronically with *P. aeruginosa* was not significant it is important to consider a subset of isolates in Cluster 1 with an evident absence of chronic PA with a high staphyloxanthin production (in concordance with isolates from mono patients), high hemolysis, variable biofilm and DNAse pattern.

Regarding the *S. aureus* background, pulsed-field electrophoresis and complemented molecular typing performed in this work showed a variety of probable *S. aureus* clonal complexes involved in CF infections, including CC1, CC5, CC45, CC22, CC30 and CC398. These lineages have been shown to cause bacteremia in Argentina and other South American countries in a recent survey, with CC398 as the most frequent of a quite diverse set of MSSA lineages and with CC5 and CC30 as the dominant MRSA clonal complexes (Di Gregorio et al., 2023; Barcudi et al., 2024). Other studies in Argentina provide results in line with these observations (Bacurdi et al., 2024). This contrasts with epidemiological data from the United States, where while more than 80% of SA belong to CC5, CC8, CC30 and CC45, and CC398 appears not to be relevant among MSSA in non-CF and CF patients (Mohamed et al., 2019; Long et al., 2021), suggesting that CF infections by *S. aureus* parallel the lineages circulating in the community. Additionally, as opposed to patients with CF in Austria showing that predominant clonal complexes were CC30 (22%), CC15 (16%), CC45 (14%), and CC5 (12%), a study carried out in Poland described the presence of CF isolates from CC30, CC22, CC97, CC45, CC15 and CC5, which was the most frequently isolated (Masoud-Landgraf et al., 2015; Garbacz et al., 2018). However, the scene appears to be more complex, as another survey found an association of CF isolates with hospital-acquired SCCmec was reported, probably indicating some role of the hospital environment (Manara et al., 2018). In a previous study, a dynamic transmission process of MRSA clones between hospitals and the community was reported (Barcudi et al., 2020), supporting the notion that circulating lineages in both settings can also colonize patients with CF. Interestingly, in our study similar genetic backgrounds were not associated with a given cluster, highlighting the possible role of the microenvironment in selective pressure. Bernardy et al (2020) showed different hemolytic activity, polysaccharide production and resistance towards mucoid and nonmucoid *P. aeruginosa* PAO1 of strains of *S. aureus* isolated from CF patients within CC 5, 8 and 30. Furthermore, Wieneke et al (2021) evaluated hemolysis, beta-toxin, mucoid, DNase activity, SCV morphotype, biofilm formation and pigment of SA isolated from sputa of CF patients and found phenotypic diversity within related spa-types.

Another complexity layer is the intrinsic complex regulation network of *S. aureus* including global regulators systems Agr, SaeSR, Sar, MgrA, CodY among others. Rumpf et al. (2021), performed an exhaustive analysis of different longitudinal studies in CF patients analyzing mutation rates, gene transference, gene expression, gene regulation and phenotypic heterogeneity. These authors remark on the importance of analyzing more than one gene to understand the *S. aureus* virulence. In this work the three clusters showed a specific phenotypic patterns that are compatible with some specific regulation landscapes, for example strains belonging to Cluster 1 could be assimilable with a higher SigB activity (leading to high staphyloxanthin production and biofilm formation, low DNAse activity and hemolysis variable); Cluster 2 could respond to a SarA and SigB conserved function and a variant of Agr with a low but not null activity (low staphyloxanthin production, low hemolysis and average biofilm and DNAse activity) and Cluster 3 respond to SigB low activity (low biofilm and high hemolysis and DNAse). In CF isolates, negative variants of *sigB, agr* and other regulator genes have been described (Rumpf et al., 2021).

Finally, we focused on two patients with similar characteristics and observed the emergence of MRSA isolates after a transitory PA infection treated with ciprofloxacin. Further *in vitro* experiments performed using one MRSA and one MSSA isolates from P2 with the same ciprofloxacin MIC showed a higher survival of the MRSA isolate over the MSSA only when the antibiotic was added in a subinhibitory concentration to co-cultures with *P. aeruginosa* PA14. In polymicrobial infections, *P. aeruginosa* PA14 exerts an antagonistic effect on *S. aureus*, driving transcriptomic remodeling of the latter. Co-culture studies have shown that PA14 induces oxidative and nitrosative stress responses in *S. aureus*, while simultaneously repressing pathways linked to energy metabolism and virulence (Tognon et al., 2017; Noto et al., 2017). Importantly, multidrug efflux pumps such as Nor family transporters are upregulated during these interactions, potentially enhancing *S. aureus* tolerance to fluoroquinolones like ciprofloxacin (Bayer et al., 2013; Kaatz et al., 2005a and b). This is consistent with our observation that MRSA isolates displayed increased survival in PA14+ciprofloxacin co-cultures, a phenotype that might be linked to NorC-mediated efflux. Together, these findings underscore how interspecies interactions can potentiate antimicrobial resilience beyond canonical resistance determinants.

All these aspects showed the complexity of the analysis, however our results showed some key points: i) SA-M showed a higher staphyloxanthin production and higher presence of non-hemolytic strains ii) despite the variability the SA isolates can be clustered in only 3 groups of phenotypic pattern with statistical relevance iii) the clustering did not depend on the ST iv) although the presence of chronic PA was not a differential factor statistically significant in Cluster 1 is evident the lack of PA in the SA with high staphyloxanthin in concordance with the higher production in isolates from monoinfected patients v) the change from one cluster to the other one seems to be driven by a change in the airway microenvironment and not due to a change in the ST vi) the presence of a transitory PA infection drives a change in the cluster of SA-T isolates from monoinfected patients vii) the change of cluster in SA-T with transitory infection seems to be mainly from Cluster 1 to 3 viii) after the transitory PA infection in 2/4 analyzed patients presented a significative higher proportion of MRSA ix) in vitro assays showed a higher survival of a MRSA isolate when is cocultured with a MSSA isolate in presence of PA14 and ciprofloxacin.

In conclusion, this longitudinal prospective study provided an integrated analysis of the dynamics of *S. aureus* infections, highlighting that transient *P. aeruginosa* infections can act as modulators of *S. aureus* phenotypic patterns, particularly following P*. aeruginosa* eradication with ciprofloxacin. Notably, in 2 out of 5 monoinfected patients, we observed a shift from MSSA to MRSA, raising concerns about the potential selection of resistant clones previously present at low abundance or the acquisition of new resistant strains. Although based on a limited number of cases, this finding along with our in vitro results underline the need to monitor such transitions more systematically to better understand their clinical relevance.

## Data Availability

All data produced in the present study are available upon reasonable request to the authors

## Acknowledgments

We thank Dr. Sergio Nemirovsky for his invaluable advice in the statistical treatment of the data.

## Financial Statement

this article was supported by “Redes Federales de Alto Impacto, Red REPARA” program (Jefatura de Gabinete, Gobierno Nacional, Argentina) to AV, AS, JG and PMT, the National Agency for Scientific and Technological Promotion PICT-2021-GRFTI-00265 and +4I program FCEyN-UBA to PT and by the National Council for Scientific Research and Technology of Argentina CONICET-PIP-2022) and the National Agency for Scientific and Technological Promotion (ANPCyT–PICT 2020-02171) to CS. JG, AS, AV, CS and PMT, SAR, MBG and MDA hold a doctoral fellowship from CONICET, EB and AF hold a ANPCyT doctoral fellowship.

