## Supplemental material for "Microenvironment-driven phenotypic shifts in *Staphylococcus aureus* from children with cystic fibrosis: a longitudinal cohort study"

### Supplementary Figures

**Supplementary Fig.1** Principal component analysis based on the mean values of the different phenotypes. A. Dimensions 1 and 2. B. Dimensions 1 and 3.

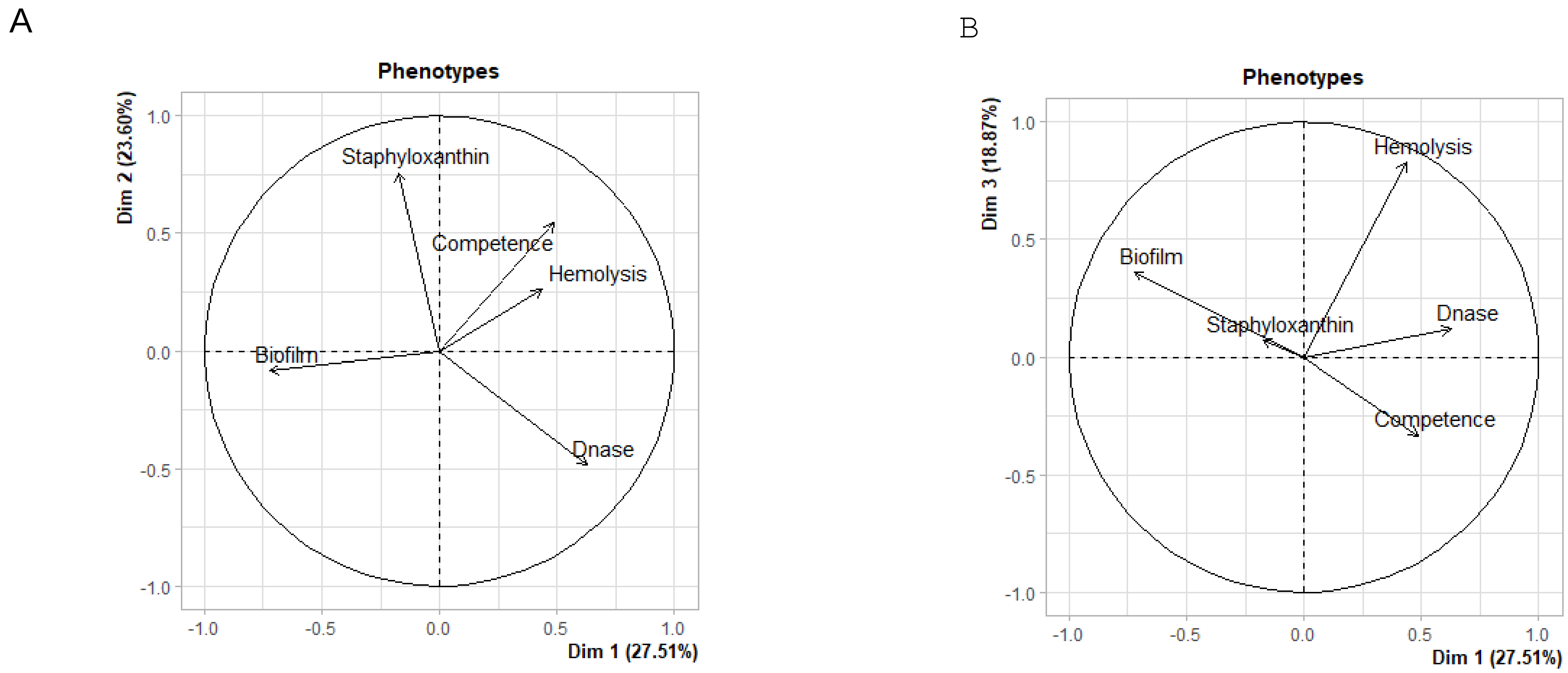

### Supplementary Figure 2

A. Patient contribution to the different clusters

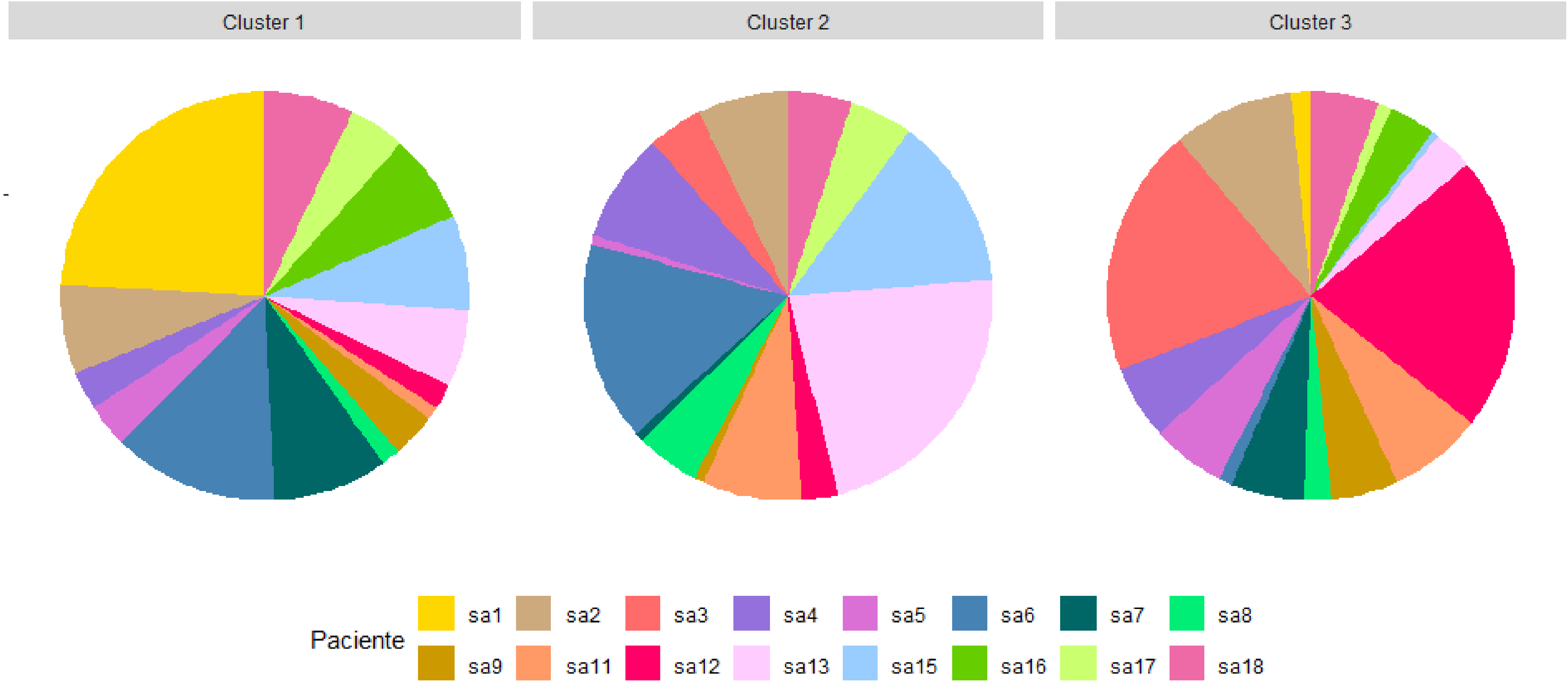

**Supplementary Figure 2**

B. SA isolate contribution to the different clusters

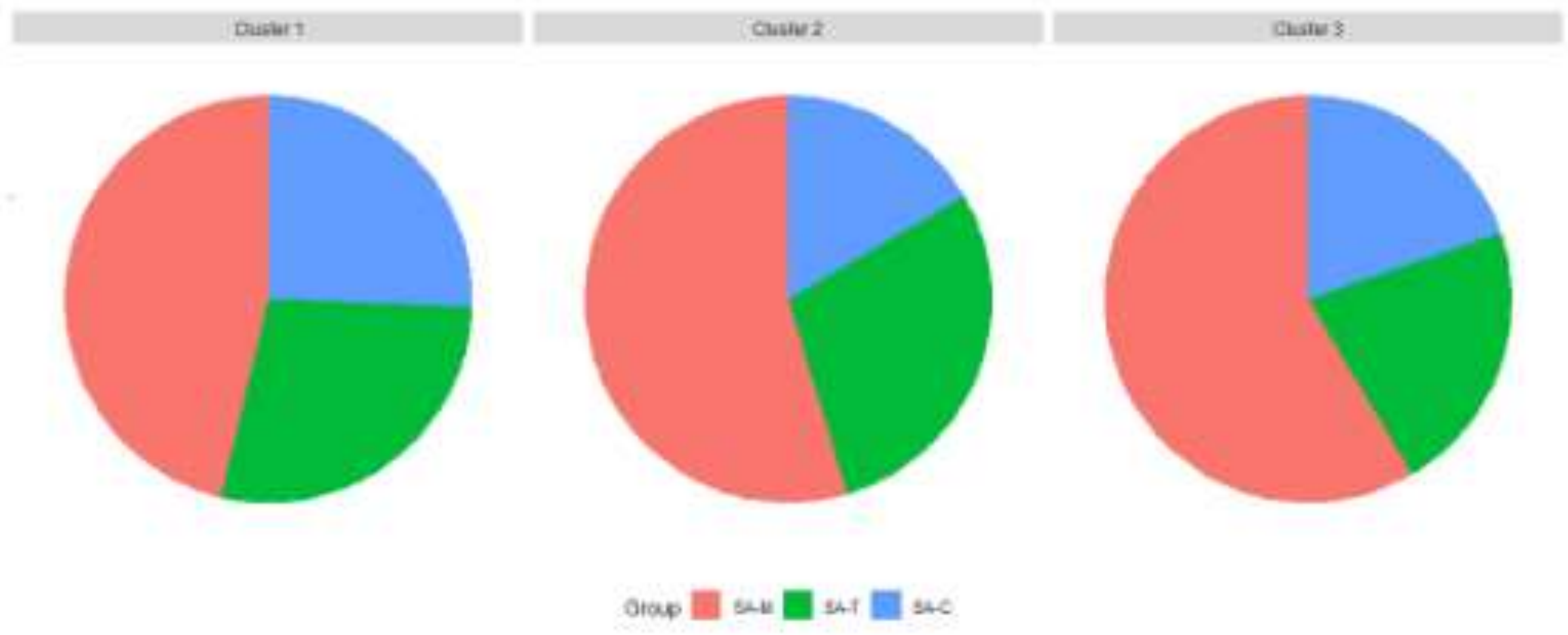

**Supplementary Figure 2**

C. Modulator contribution to the different cluster

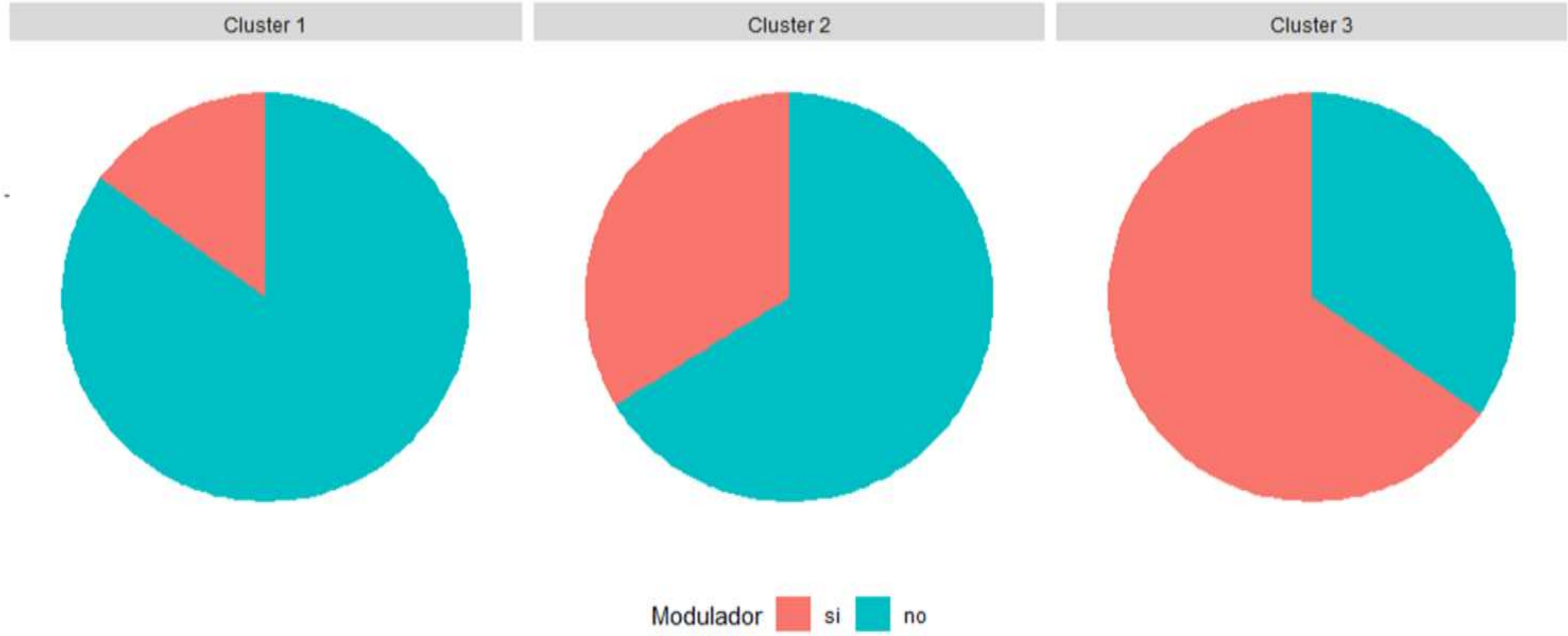

Supplementary Figure 2

C. Chronic PA contribution to the different clusters

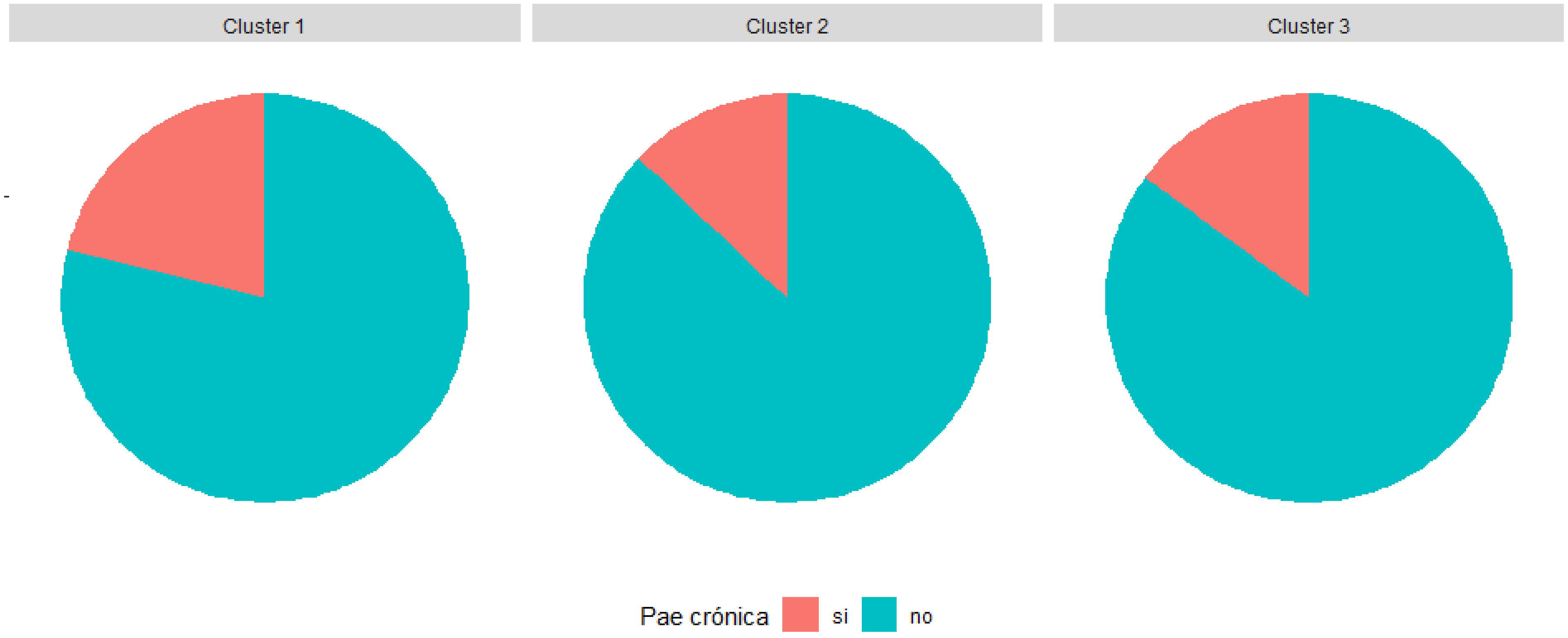

Supplementary Figure 3.A

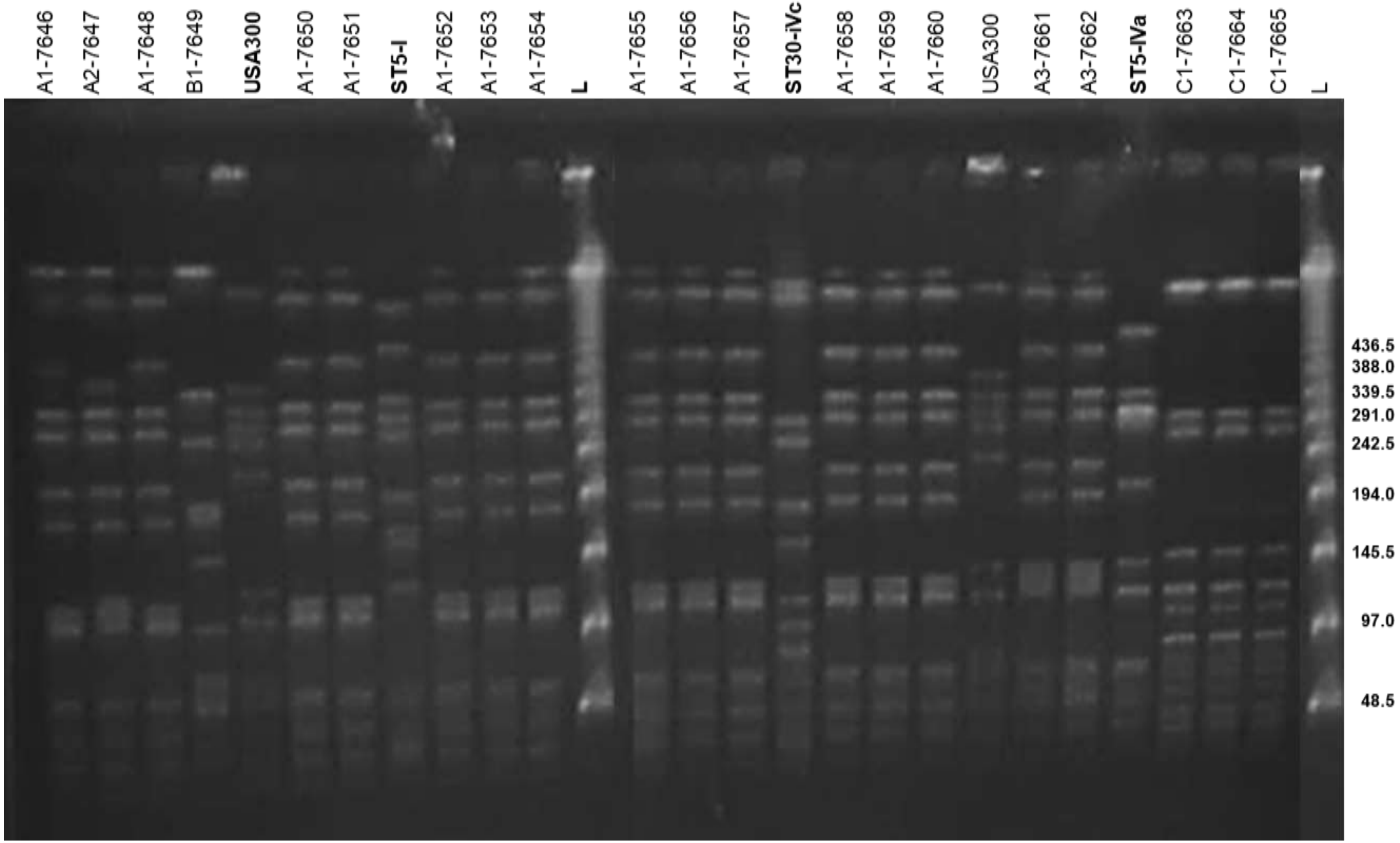


**Figure S3A**. PFGE of *S. aureus* SmaI-digested genomic DNA from selected strains recovered from patient 1. Representative isolates of ST8-IVa (USA300), ST5-I (Cordobés/Chilean clone), ST30-IVc (Southwest Pacific-SWP/USA1100 clone), and ST5-IVa (CA-MRSA clone in Argentina, similar to USA800) were included for comparison. The corresponding subtypes and strain IDs are indicated. L: DNA molecular size marker (lambda DNA ladder, Promega). NCTC 8325 was used as the control strain.

Supplementary Figure 3.B

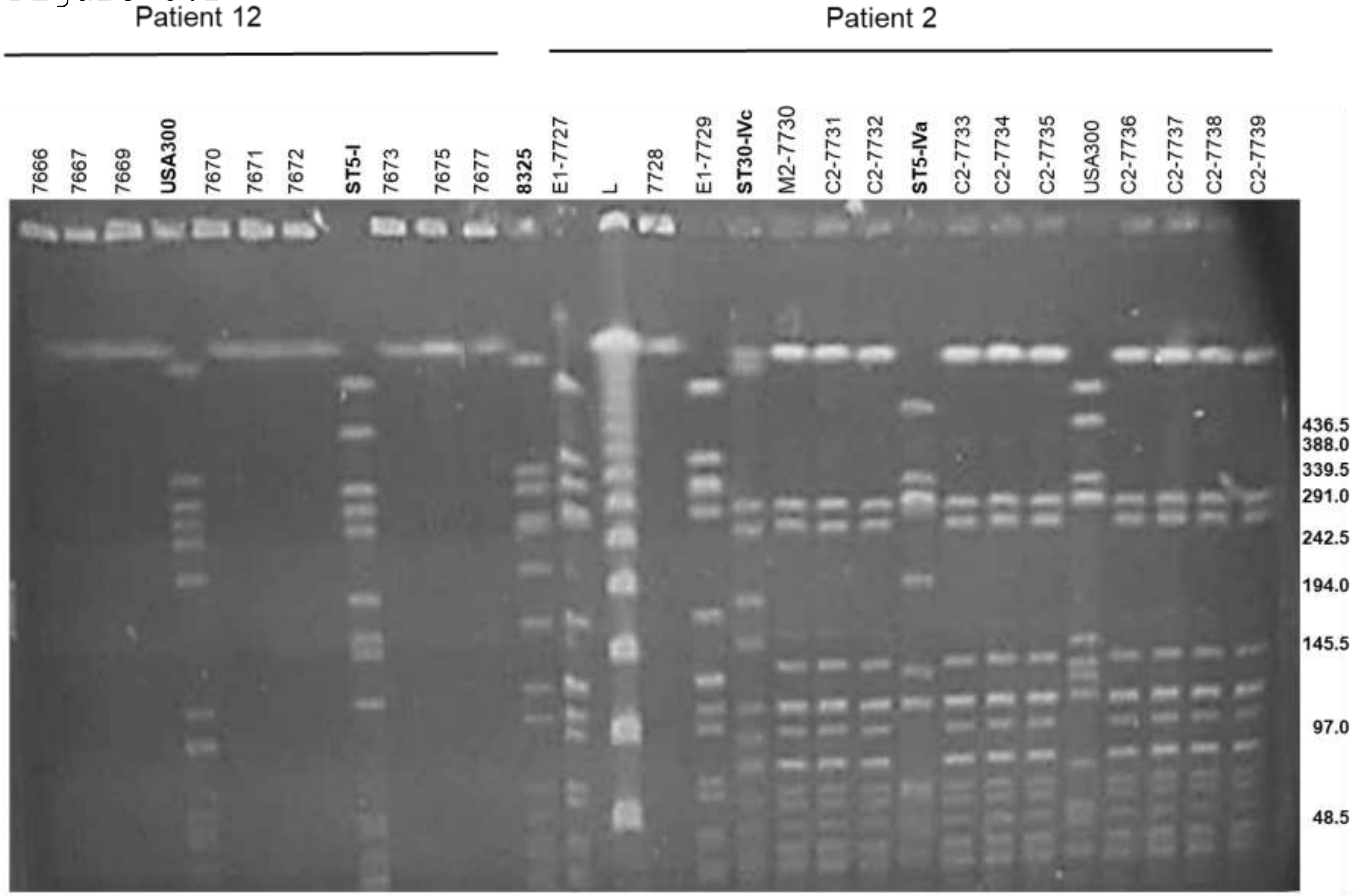


**Figure S3B**. PFGE of *S. aureus* SmaI-digested genomic DNA from selected strains recovered from patients 2 and 12. Representative isolates of ST8-IVa (USA300), ST5-I (Cordobés/Chilean clone), ST30-IVc (Southwest Pacific-SWP/USA1100 clone), and ST5-IVa (CA-MRSA clone in Argentina, similar to USA800) were included for comparison. The corresponding subtypes and strain IDs are indicated. L: DNA molecular size marker (lambda DNA ladder, Promega). NCTC 8325 was used as the control strain.

Supplementary Figure 4

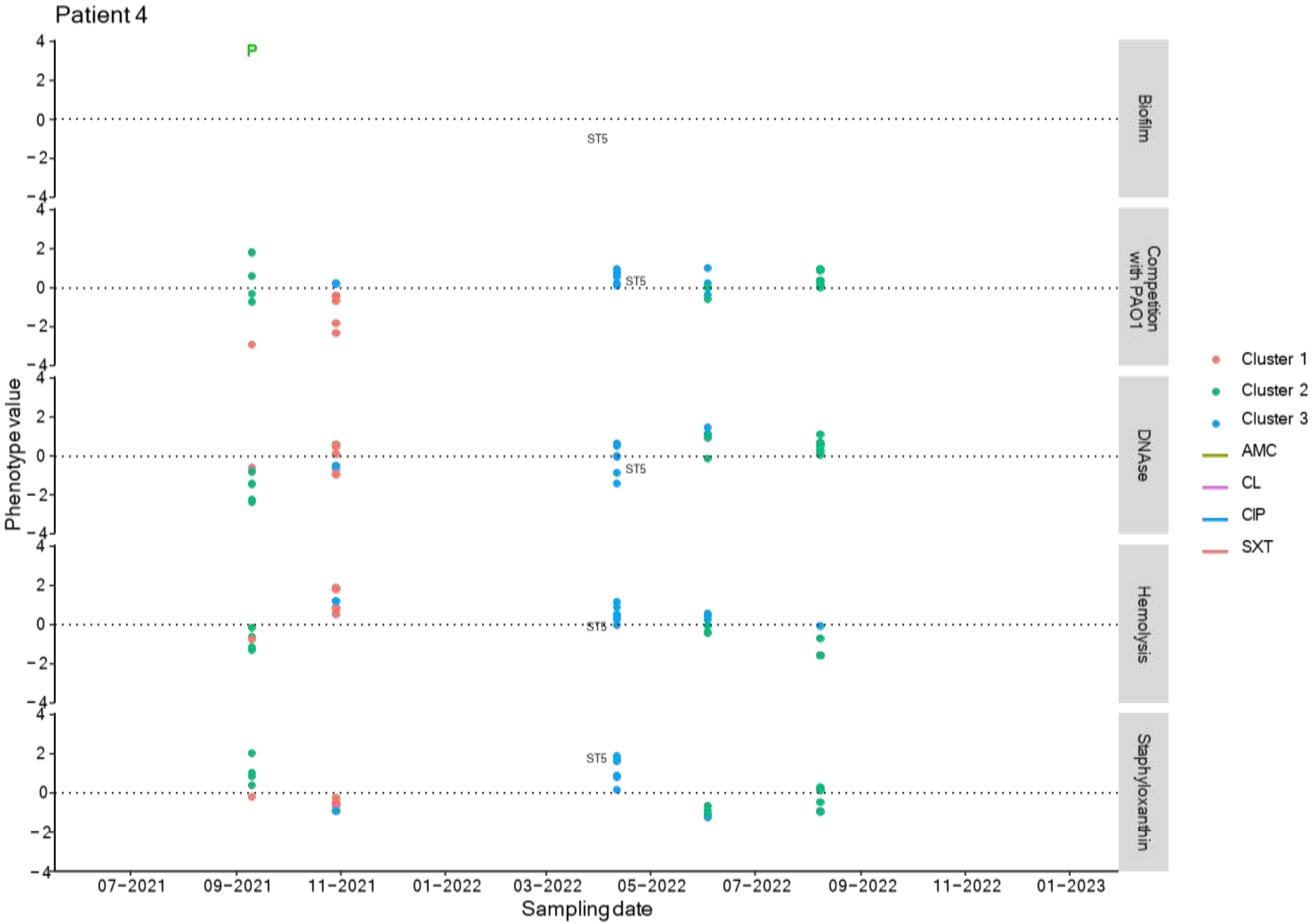

Supplementary Figure 5

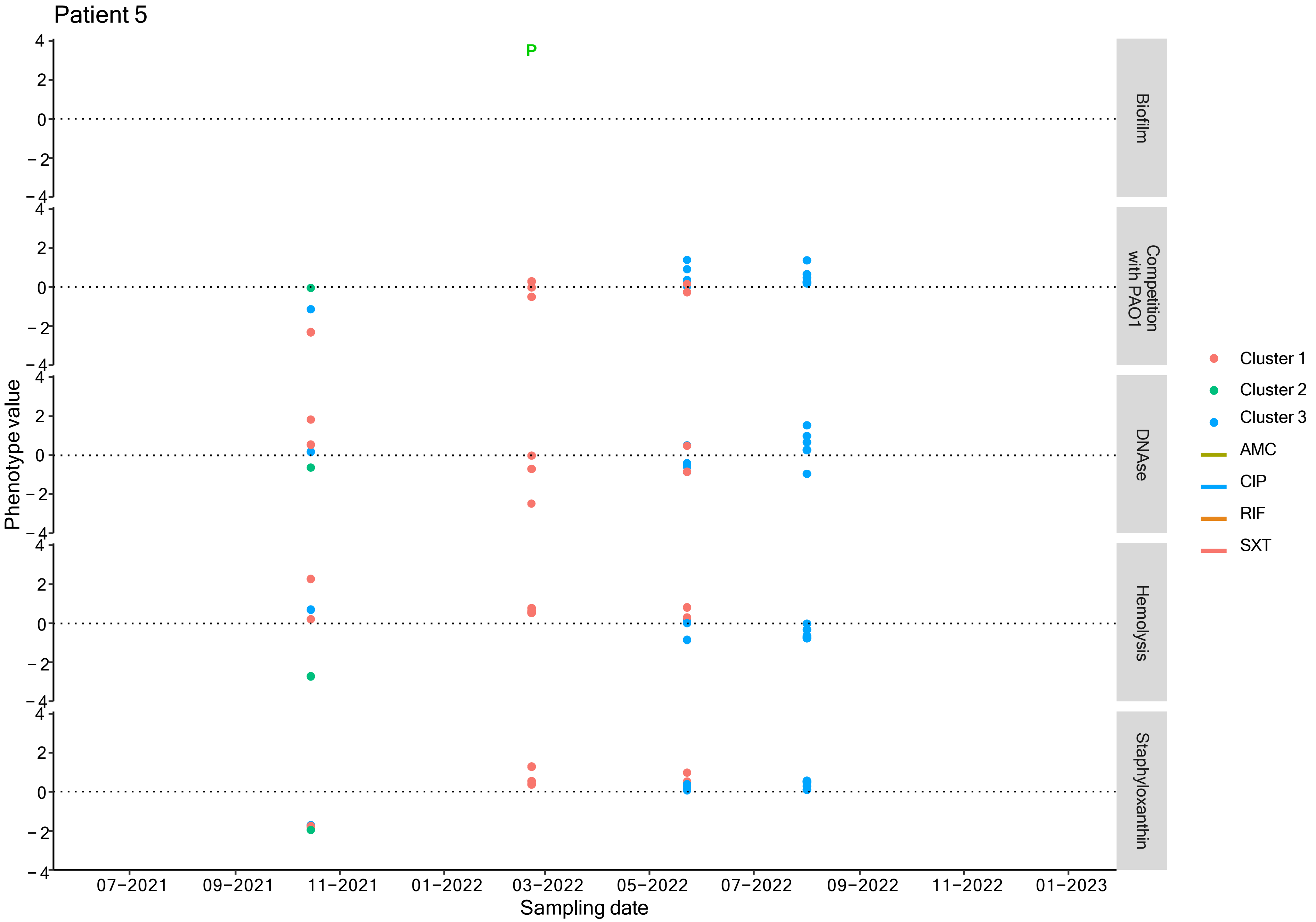

### Supplementary Figure 6

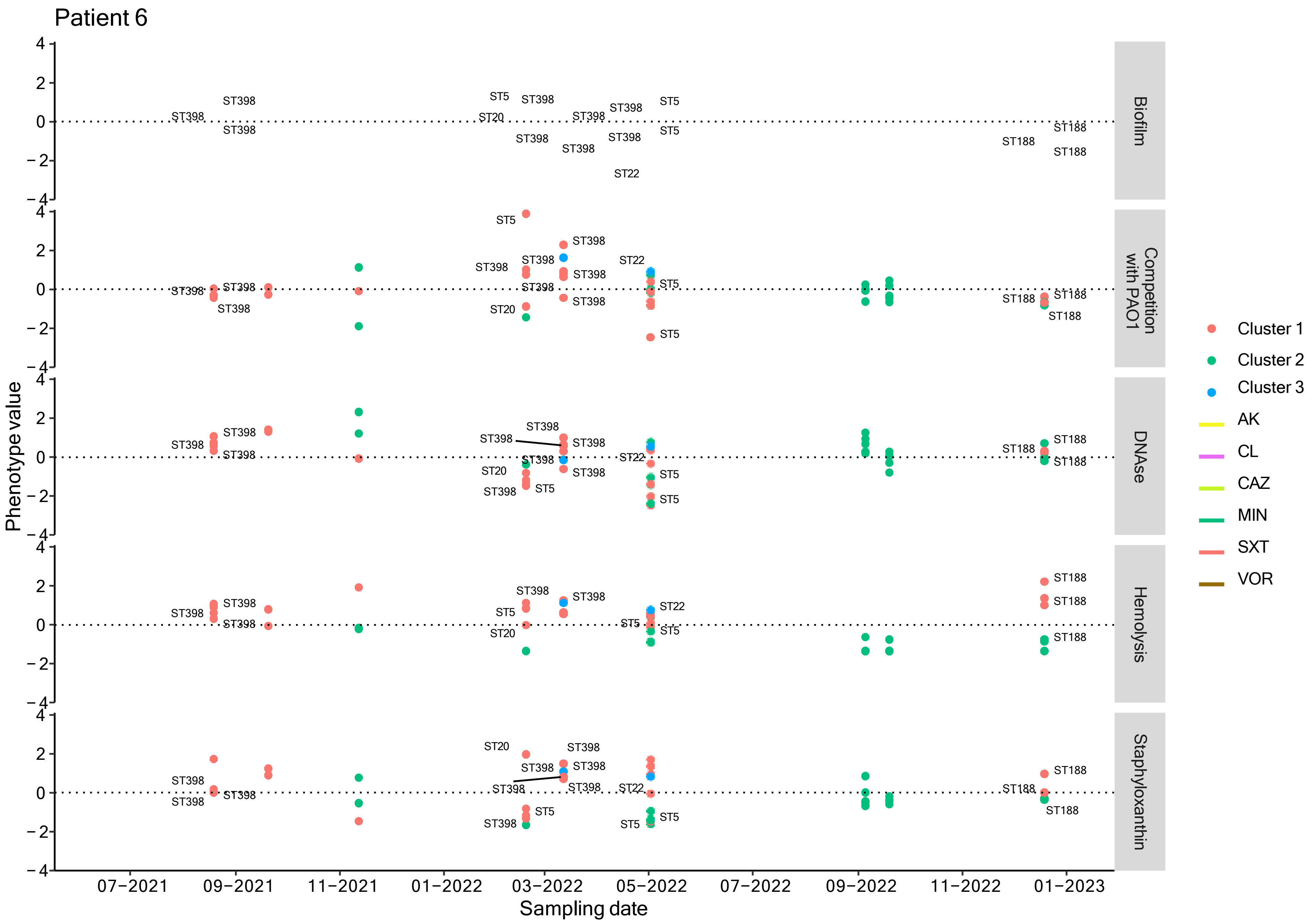

Supplementary Figure 7

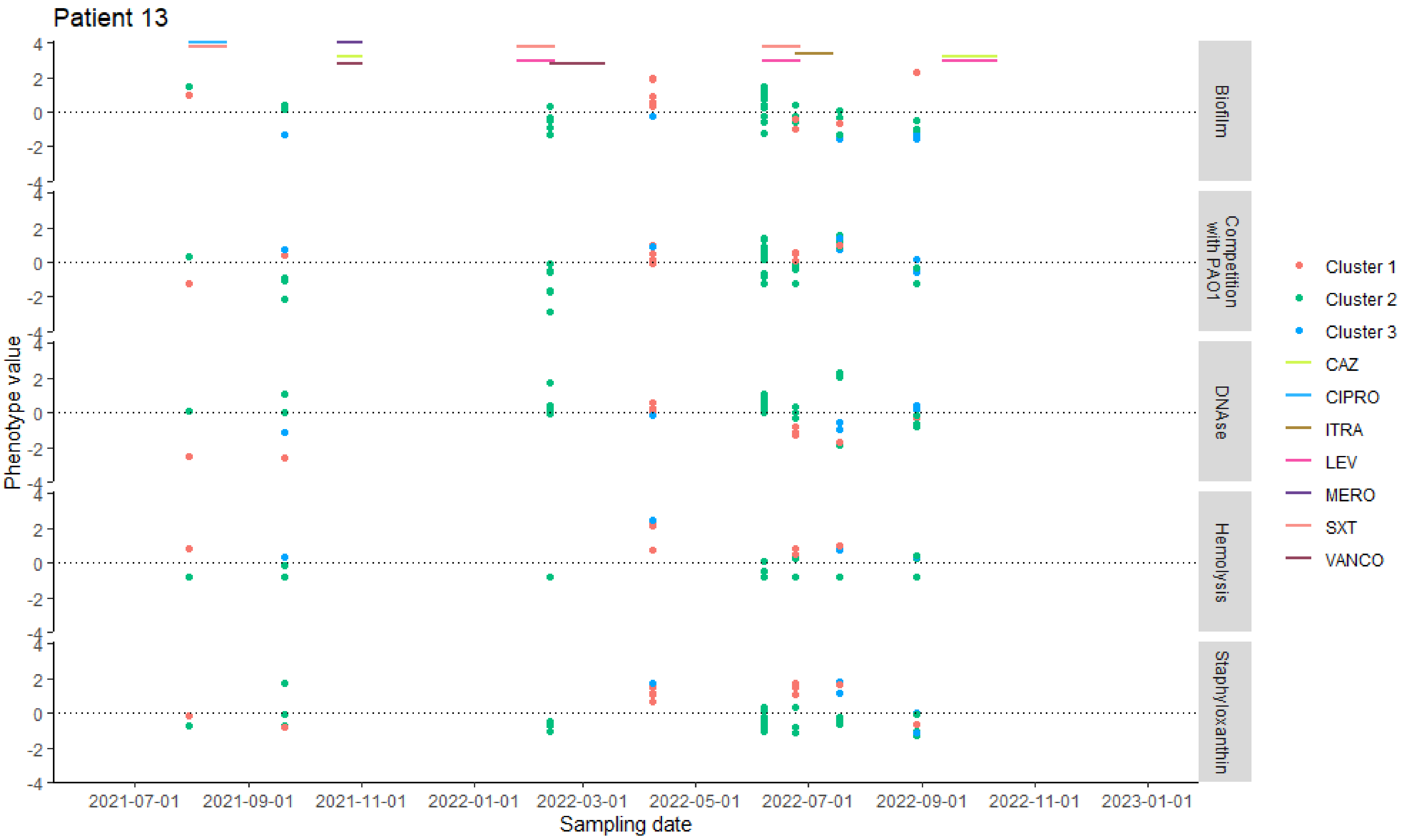

Supplementary Figure 8

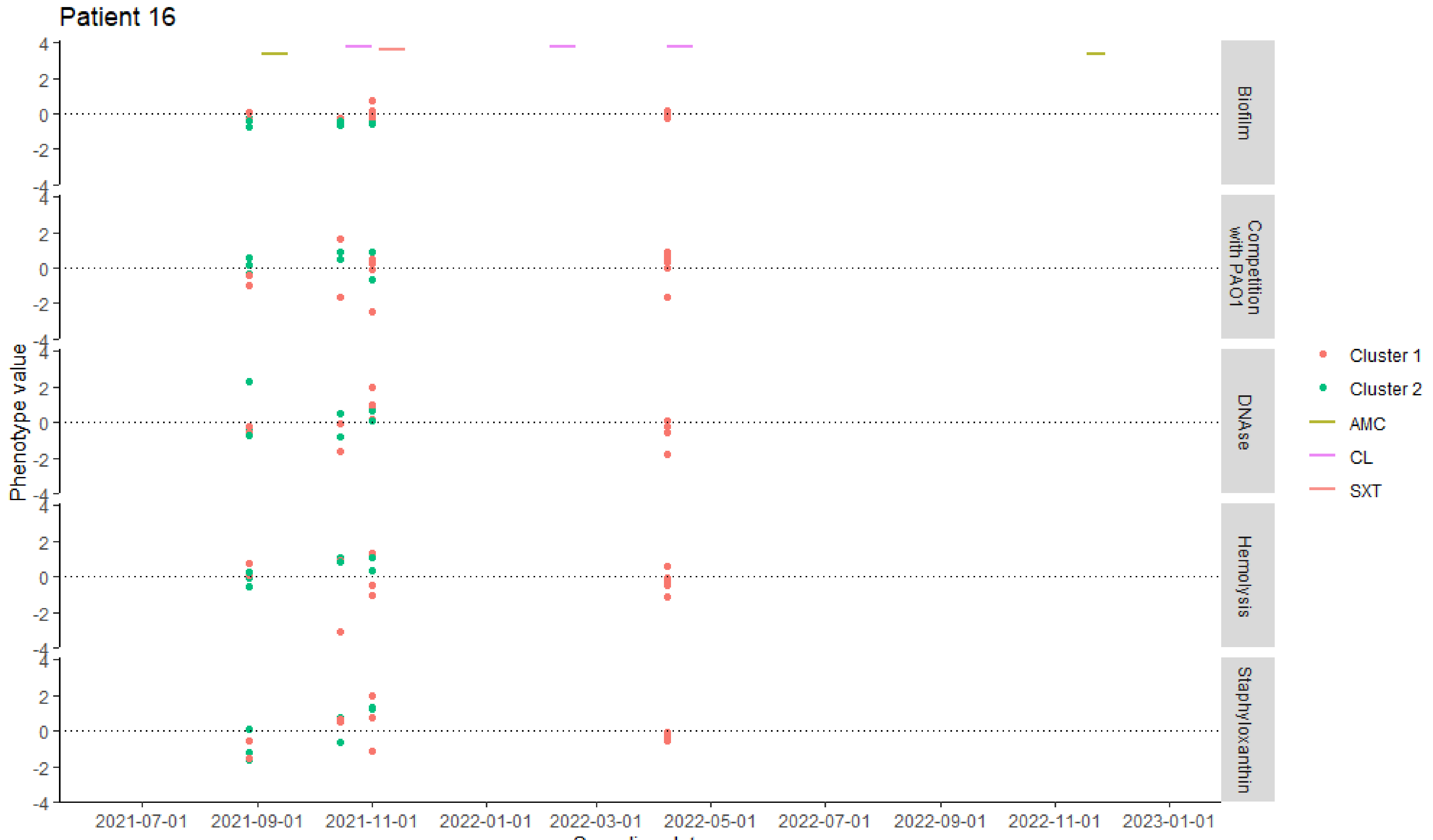

Supplementary Figure 9

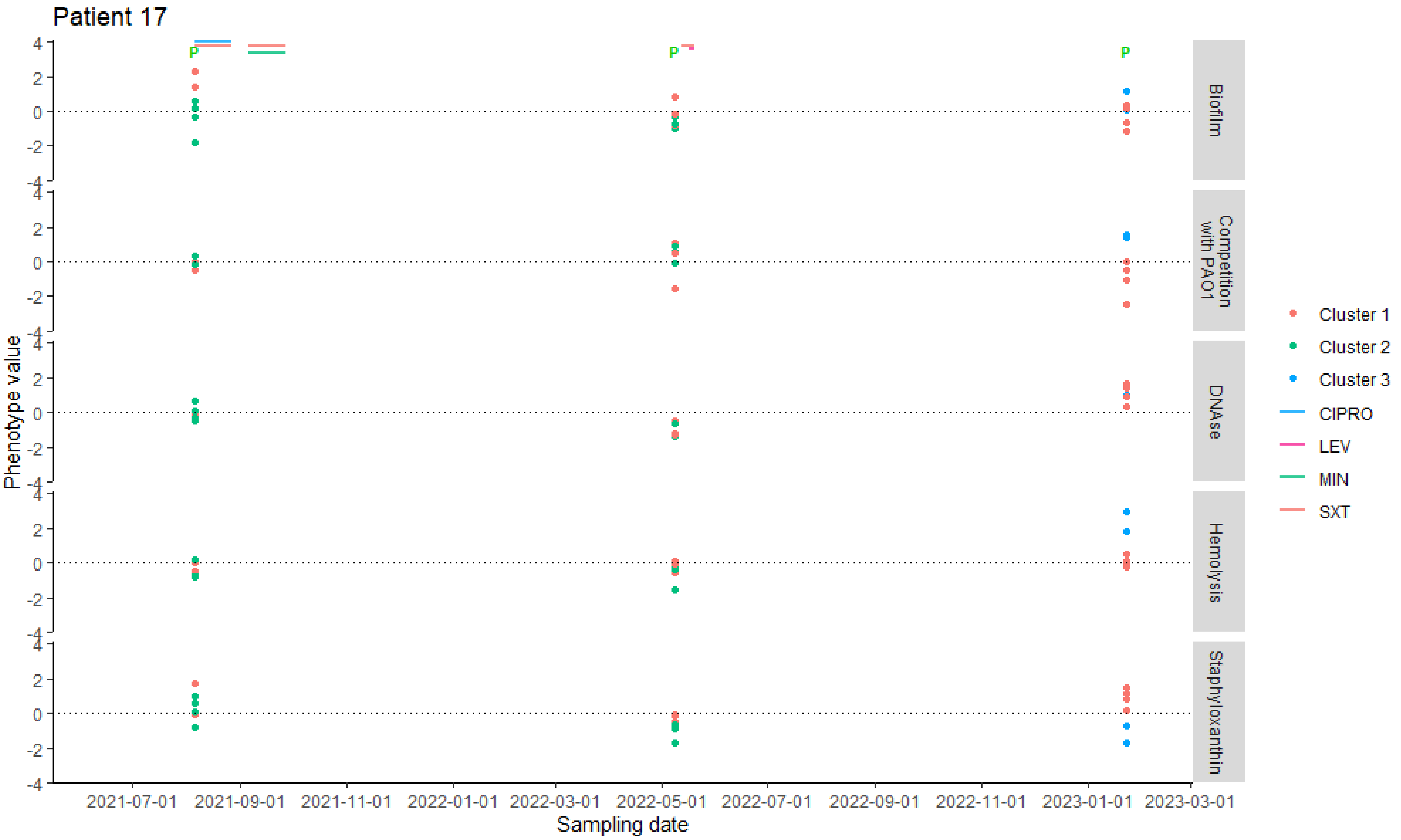

Supplementary Figure 10

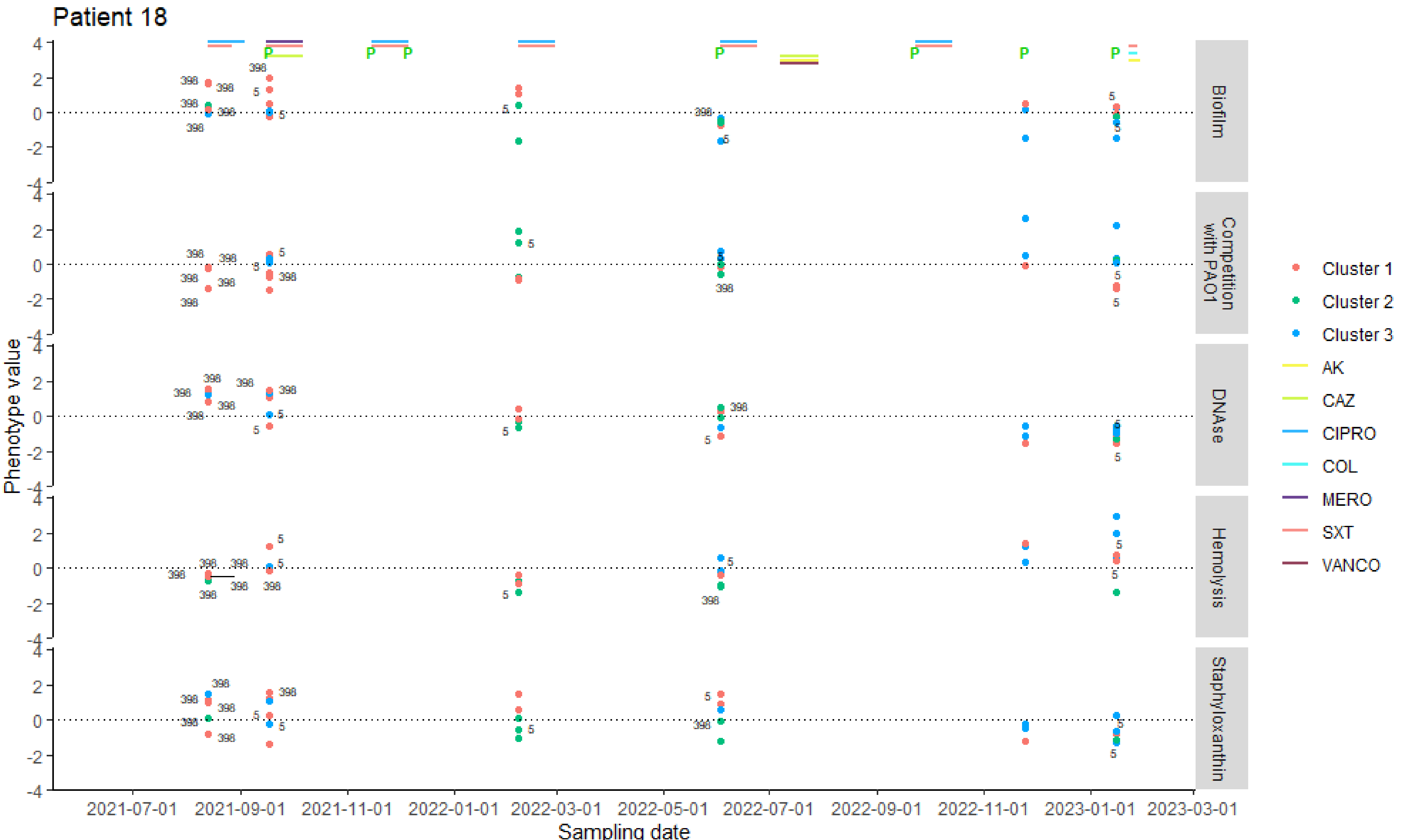

Table S1. Survival assays in presence of PA and ciprofloxacin.

| Isolate (*S. aureus*) | Co-culture with PA14 | Ciprofloxacin 0.250 µg/ml | Inoculum of S. aureus (CFU/ml) | Inoculum of *P. aeruginosa* (CFU/ml) | Ratio (SA:PA) |
|---|---|---|---|---|---|
| MSSA | Yes | No | $1x10^8$ | NA | NA |
| MRSA | Yes | No | $1x10^8$ | NA | NA |
| MRSA + MSSA | Yes | No | $1x10^8$ | NA | NA |
| MSSA | Yes | Yes | $1x10^8$ | $1x10^8$ | 1:1 |
| MRSA | Yes | Yes | $1x10^8$ | $1x10^8$ | 1:1 |
| MRSA + MSSA | Yes | Yes | $1x10^8$ | $1x10^8$ | 1:1:1 |
| MSSA | No | Yes | $1x10^8$ | NA | NA |
| MRSA | No | Yes | $1x10^8$ | NA | NA |
| MRSA + MSSA | No | Yes | $1x10^8$ | NA | NA |
| MSSA | No | No | $1x10^8$ | NA | NA |
| MRSA | No | No | $1x10^8$ | NA | NA |
| MRSA + MSSA | No | No | $1x10^8$ | NA | NA |

NA: Not applied

**Table S2: Patient characteristics**

| Variable | Value |
|---|---|
| Age (years old, interquartile range) | 8.8 (6.5 - 13.2) |
| Sex (% male) | 62.5% |
| **CFTR variant (n)** | |
| F508del heterozygotes | 8 |
| F508del homozygotes | 5 |
| Other variants | 7 |
| Other variants in absence of F508del | 2 |
| **$FEV_1$ impairment (n)** | |
| Mild | 7 |
| Moderate | 3 |
| Severe | 3 |
| Spirometry technique not achieved | 3 |
| **Clinical course** | |
| Total pulmonary exacerbations (median, range) | 6 (1-9) |
| Pulmonary exacerbations requiring hospitalization (median, range) | 0 (0-5) |
| **Pulmonary therapies** | |
| Hypertonic solution (%) | 81 |
| DNAse (%) | 81 |
| Macrolides (%) | 38 |
| Supplementary oxygen (%) | 19 |
| **CFTR modulators** | |

| | |
|---|---|
| Receiving modulator therapy at the onset of sampling | 2 |
| Receiving modulator therapy at the end of sampling | 10 |
| Lumacaftor/ivacaftor | 2 |
| Elexacaftor/tezacaftor/ivacaftor | 8 |
| **Inhaled antibiotics** | |
| Tobramycin (%) | 69 |
| Colistin (%) | 31 |
| Vancomycin (%) | 13 |
| **Microbiological studies** | |
| Cultures performed (median, range) | 9.5 (5-16) |
| *Staphylococcus aureus* isolates collected (median, IQR) | 30.5 (20.5-44.5) |
| Chronic infection with *Staphylococcus aureus* (%) | 100 |
| Chronic infection with *Pseudomonas aeruginosa* (%) | 31 |

**Table S3.** Correlation between phenotypes using Spearman's rank

| Phenotype | Staphyloxantin | Hemolysis | Competence | Biofilm | DNAse |
|---|---|---|---|---|---|
| **Staphyloxantin** | - | | | | -0.15, $p<0.001$ |
| **Hemolysis** | | - | | | 0.13, $p<0.001$ |
| **Competence** | | | - | 0.13, $p < 0.001$ | 0.21, $p<0.001$ |
| **Biofilm** | | | 0.13, $p < 0.001$ | - | -0.23, $p < 0.001$ |
| **DNAse** | -0.15, $p<0.001$ | 0.13, $p<0.001$ | 0.21, $p<0.001$ | -0.23, $p < 0.001$ | - |

**Table S4. PCA score table based on the mean values of the different phenotypes**

#### Eigenvalues and variance explained

| Component | Eigenvalue | % of variance | Cumulative % of variance |
|---|---|---|---|
| Comp 1 | 1.3757011 | 27.514 | 27.514 |
| Comp 2 | 1.1802252 | 23.605 | 51.119 |
| Comp 3 | 0.9435452 | 18.871 | 69.989 |
| Comp 4 | 0.8219584 | 16.439 | 86.429 |
| Comp 5 | 0.6785701 | 13.571 | 100.000 |

#### Coordinates

| Variable | Dim.1 | Dim.2 | Dim.3 | Dim.4 | Dim.5 |
|---|---|---|---|---|---|
| Mean staphyloxanthin | -0.1753919 | 0.7585286 | 0.0717161 | -0.5856725 | 0.2138143 |
| log(mean biofilm) | -0.7234283 | -0.0839758 | 0.3601417 | 0.3103622 | 0.4935309 |
| log(mean competence) | 0.4869344 | 0.5430106 | -0.3326841 | 0.5071358 | 0.3164949 |
| log(mean hemolysis) | 0.435404 | 0.2593978 | 0.8268902 | 0.1649415 | -0.1793962 |
| log(mean DNase) | 0.6284175 | -0.4854482 | 0.1194733 | -0.313415 | 0.5068814 |

#### Correlation

| Variable | Dim.1 | Dim.2 | Dim.3 | Dim.4 | Dim.5 |
|---|---|---|---|---|---|
| Mean staphyloxanthin | -0.1753919 | 0.7585286 | 0.0717161 | -0.5856725 | 0.2138143 |
| log(mean biofilm) | -0.7234283 | -0.0839758 | 0.3601417 | 0.3103622 | 0.4935309 |
| log(mean competence) | 0.4869344 | 0.5430106 | -0.3326841 | 0.5071358 | 0.3164949 |
| log(mean hemolysis) | 0.435404 | 0.2593978 | 0.8268902 | 0.1649415 | -0.1793962 |
| log(mean DNase) | 0.6284175 | -0.4854482 | 0.1194733 | -0.313415 | 0.5068814 |

#### Squared cosine ($cos^2$)

| Variable | Dim.1 | Dim.2 | Dim.3 | Dim.4 | Dim.5 |
|---|---|---|---|---|---|
| Mean staphyloxanthin | 0.0307623 | 0.5753656 | 0.0051432 | 0.3430123 | 0.0457166 |

| | | | | | |
|---|---|---|---|---|---|
| log(mean biofilm) | 0.5233486 | 0.0070519 | 0.1297020 | 0.0963247 | 0.2435728 |
| log(mean competence) | 0.2371051 | 0.2948605 | 0.1106787 | 0.2571867 | 0.1001690 |
| log(mean hemolysis) | 0.1895767 | 0.0672872 | 0.6837474 | 0.0272057 | 0.0321830 |
| log(mean DNase) | 0.3949085 | 0.2356599 | 0.0142739 | 0.0982289 | 0.2569288 |

#### Contribution (%)

| Variable | Dim.1 | Dim.2 | Dim.3 | Dim.4 | Dim.5 |
|---|---|---|---|---|---|
| Mean staphyloxanthin | 2.2361 | 48.7505 | 0.5451 | 41.7311 | 6.7372 |
| log(mean biofilm) | 38.0423 | 0.5975 | 13.7462 | 11.7189 | 35.8950 |
| log(mean competence) | 17.2352 | 24.9834 | 11.7301 | 31.2895 | 14.7618 |
| log(mean hemolysis) | 13.7804 | 5.7012 | 72.4658 | 3.3099 | 4.7428 |
| log(mean DNase) | 28.7060 | 19.9674 | 1.5128 | 11.9506 | 37.8633 |

**Table S5:** Molecular characterization of selected SA isolates

| Isolates with PFGE | Patient | PFGE type | RIDOM *spa* type | CC398/*sau1-hsdS1* | Sequence Type (ST) | Cluster | Sampling Date |
|---|---|---|---|---|---|---|---|
| AW-7646 | P1 | A1 | t1987 | neg | ST20 | 1 | Jul-21 |
| AV-7647 | P1 | A2 | t1987 | neg | ST20 | 1 | Jul-21 |
| AU-7648 | P1 | A1 | | neg | ST20 | 1 | Jul-21 |
| AT-7649 | P1 | B1 | NA/09-02-16-34-13-17-34-23-34-34 | neg | ST45 | 1 | Jul-21 |
| HM-7650 | P1 | A1 | | neg | ST20 | 1 | Mar-22 |
| HN-7651 | P1 | A1 | t1987 | neg | ST20 | 1 | Mar-22 |
| HÑ-7652 | P1 | A1 | | neg | ST20 | 1 | Mar-22 |
| HO-7653 | P1 | A1 | | neg | ST20 | 1 | Mar-22 |
| HP-7654 | P1 | A1 | t1987 | neg | ST20 | 1 | Mar-22 |
| HR-7655 | P1 | A1 | | neg | ST20 | 1 | May-22 |
| HU-7656 | P1 | A1 | | neg | ST20 | 1 | May-22 |
| HQ-7657 | P1 | A1 | t1987 | neg | ST20 | 1 | May-22 |
| PH-7658 | P1 | A1 | | neg | ST20 | 3 | Aug-22 |
| PI-7659 | P1 | A1 | t1987 | neg | ST20 | 1 | Aug-22 |
| PJ-7660 | P1 | A1 | | neg | ST20 | 1 | Aug-22 |
| SU-7661 | P1 | A3 | t1987 | neg | ST20 | 1 | Dec-22 |
| SW-7662 | P1 | A3 | | neg | ST20 | 1 | Dec-22 |
| UB-7663 | P1 | C1 | t665 | neg | ST30 | 1 | Dec-22 |
| UA-7664 | P1 | C1 | | neg | ST30 | 1 | Dec-22 |
| TY-7665 | P1 | C1 | | neg | ST30 | 1 | Dec-22 |
| Y-7666 | P12 | NT | t1451 | pos | ST398 | 1 | Aug-21 |
| Z-7667 | P12 | NT | | pos | ST398 | 3 | Aug-21 |
| FE-7669 | P12 | NT | | pos | ST398 | 3 | Feb-22 |
| FF-7670 | P12 | NT | | pos | ST398 | 3 | Feb-22 |
| FH-7671 | P12 | NT | | pos | ST398 | 3 | Feb-22 |
| FG-7672 | P12 | NT | | pos | ST398 | 1 | Feb-22 |
| PL-7673 | P12 | NT | | pos | ST398 | 3 | Aug-22 |
| PM-7674 | P12 | NT | | pos | ST398 | 3 | Aug-22 |
| PN-7675 | P12 | NT | t1451 | pos | ST398 | 3 | Aug-22 |
| PP-7677 | P12 | NT | | pos | ST398 | 3 | Aug-22 |
| AO-7678 | P15 | D1 | t002 | neg | ST5 | 1 | Aug-21 |
| AP-7679 | P15 | D1 | | neg | ST5 | 1 | Aug-21 |
| CA-7680 | P15 | D1 | | neg | ST5 | 1 | Nov-21 |
| CE-7681 | P15 | D1 | t002 | neg | ST5 | 1 | Nov-21 |
| CC-7682 | P15 | D1 | | neg | ST5 | 1 | Nov-21 |
| JS-7683 | P15 | D1 | | neg | ST5 | 2 | Apr-22 |
| JT-7684 | P15 | D1 | | neg | ST5 | 2 | Apr-22 |
| JV-7685 | P15 | D1 | | neg | ST5 | 3 | Apr-22 |
| LL-7686 | P15 | D1 | | neg | ST5 | 2 | Jun-22 |
| LO-7687 | P15 | D1 | | neg | ST5 | 2 | Jun-22 |

| | | | | | | | |
|---|---|---|---|---|---|---|---|
| LK-7688 | P15 | D1 | | neg | ST5 | 2 | Jun-22 |
| RA-7689 | P15 | D1 | | neg | ST5 | 2 | Sep-22 |
| RE-7690 | P15 | D1 | | neg | ST5 | 2 | Sep-22 |
| RB-7691 | P15 | D1 | | neg | ST5 | 2 | Sep-22 |
| RC-7692 | P15 | D1 | | neg | ST5 | 2 | Sep-22 |
| RD-7693 | P15 | D1 | | neg | ST5 | 2 | Sep-22 |
| EJ-7694 | P18 | NT | t1451 | pos | ST398 | 2 | Aug-21 |
| EK-7695 | P18 | NT | | pos | ST398 | 1 | Aug-21 |
| EL-7696 | P18 | NT | | pos | ST398 | 3 | Aug-21 |
| EM-7697 | P18 | NT | | pos | ST398 | 1 | Aug-21 |
| EN-7698 | P18 | NT | | pos | ST398 | 1 | Aug-21 |
| ED-7699 | P18 | D2 | t002 | neg | ST5 | 1 | Sep-21 |
| EF-7700 | P18 | D2 | | neg | ST5 | 3 | Sep-21 |
| EI-7701 | P18 | NT | t1451 | pos | ST398 | 1 | Sep-21 |
| FM-7703 | P18 | D2 | | neg | ST5 | 2 | Feb-22 |
| MS-7705 | P18 | D2 | | neg | ST5 | 1 | Jun-22 |
| MV-7706 | P18 | NT | | POS | ST398 | 2 | Jun-22 |
| UL-7708 | P18 | D2 | | neg | ST5 | 1 | Jan-23 |
| UN-7709 | P18 | D2 | | neg | ST5 | 1 | Jan-23 |
| GT-7710 | P9 | B3 | t065 | neg | ST45 | 1 | Mar-22 |
| GU-7711 | P9 | B2 | | neg | ST45 | 1 | Mar-22 |
| GW-7712 | P9 | B3 | | neg | ST45 | 1 | Mar-22 |
| GY-7713 | P9 | B2 | | neg | ST45 | 1 | Mar-22 |
| GZ-7714 | P9 | B2 | t065 | neg | ST45 | 1 | Mar-22 |
| MN-7715 | P9 | NT | | pos | ST398 | 3 | May-22 |
| MÑ-7716 | P9 | NT | | pos | ST398 | 3 | May-22 |
| MO-7717 | P9 | NT | | pos | ST398 | 2 | May-22 |
| MP-7718 | P9 | NT | t1451 | pos | ST398 | 3 | May-22 |
| MQ-7719 | P9 | NT | t1451 | pos | ST398 | 3 | May-22 |
| MR-7720 | P9 | NT | | pos | ST398 | 3 | May-22 |
| OZ-7721 | P9 | NT | | pos | ST398 | 3 | Aug-22 |
| PA-7722 | P9 | NT | t1451 | pos | ST398 | 3 | Aug-22 |
| PB-7723 | P9 | NT | | pos | ST398 | 3 | Aug-22 |
| PC-7724 | P9 | NT | | pos | ST398 | 1 | Aug-22 |
| PD-7725 | P9 | NT | t1451 | pos | ST398 | 3 | Aug-22 |
| PE-7726 | P9 | NT | | pos | ST398 | 3 | Aug-22 |
| DW-7727 | P2 | E1 | NA: 11-10-21-17-34-24-34-34-24-34-34-22-25 | neg | ST8 | 1 | Sep-21 |
| DY-7728 | P2 | NT | t1184 | pos | ST398 | 1 | Sep-21 |
| EC-7729 | P2 | E1 | | neg | ST8 | 3 | Sep-21 |
| FQ-7730 | P2 | C2 | t665 | neg | ST30 | 2 | Feb-22 |
| FO-7731 | P2 | C2 | | neg | ST30 | 1 | Feb-22 |
| FT-7732 | P2 | C2 | | neg | ST30 | 1 | Feb-22 |
| NZ-7733 | P2 | C2 | | neg | ST30 | 3 | Sep-22 |

| | | | | | | | |
|---|---|---|---|---|---|---|---|
| OB-7734 | P2 | C2 | t665 | neg | ST30 | 3 | Sep-22 |
| OA-7735 | P2 | C2 | | neg | ST30 | 3 | Sep-22 |
| RQ-7736 | P2 | C2 | | neg | ST30 | 3 | Sep-22 |
| RT-7737 | P2 | C2 | | neg | ST30 | 3 | Sep-22 |
| RR-7738 | P2 | C2 | | neg | ST30 | 1 | Sep-22 |
| RS-7739 | P2 | C2 | t665 | neg | ST30 | 1 | Sep-22 |
| AH-7740 | P6 | NT | | pos | ST398 | 1 | Aug-21 |
| AI-7741 | P6 | NT | t6587 | pos | ST398 | 1 | Aug-21 |
| AF-7742 | P6 | NT | | pos | ST398 | 1 | Aug-21 |
| GC-7745 | P6 | D3 | t002 | neg | ST5 | 1 | Feb-22 |
| GG-7747 | P6 | NT | t6587 | pos | ST398 | 1 | Feb-22 |
| GI-7748 | P6 | A1 | t1987 | neg | ST20 | 1 | Feb-22 |
| HL-7749 | P6 | NT | t6587 | pos | ST398 | 1 | Mar-22 |
| HI-7750 | P6 | NT | | pos | ST398 | 1 | Mar-22 |
| HG-7751 | P6 | NT | t6587 | pos | ST398 | 1 | Mar-22 |
| HH-7752 | P6 | NT | | pos | ST398 | 1 | Mar-22 |
| HK-7753 | P6 | NT | | pos | ST398 | 1 | Mar-22 |
| IV-7754 | P6 | D4 | t4255 | neg | ST5 | 1 | May-22 |
| IT-7755 | P6 | D4 | | neg | ST5 | 1 | May-22 |
| JC-7756 | P6 | EMRSA-15 | t223 | neg | ST22 | 3 | May-22 |
| TS-7757 | P6 | G1 | t189 | neg | ST188 | 1 | Dec-22 |
| TV-7758 | P6 | G1 | t189 | neg | ST188 | 2 | Dec-22 |
| TX-7759 | P6 | G1 | t189 | neg | ST188 | 1 | Dec-22 |

NT: PFGE non-typeable strain
